# Phylogenomics, Epigenomics, Virulome, and Mobilome of Gram-negative Bacteria Co-resistant to Carbapenems and Polymyxins: A One-Health Systematic Review and Meta-analyses

**DOI:** 10.1101/2021.07.03.21259964

**Authors:** Winnie Thabisa Ramaloko, John Osei Sekyere

**Affiliations:** Department of Medical Microbiology, School of Medicine, Faculty of Health Sciences, University of Pretoria, South Africa.

**Keywords:** Carbapenem, Polymyxins, One Health, Resistome, resistance plasmids, methylome, virulome

## Abstract

Gram-negative bacteria (GNB) continue to develop resistance against important antibiotics including last-resort ones such as carbapenems and polymyxins. An analysis of GNB with co-resistance to carbapenems and polymyxins from a One Health perspective is presented.

Data of species name, country, source of isolation, resistance genes (ARGs), plasmid type, clones, and mobile genetic elements (MGEs) were deduced from 129 articles from January 2016 to March 2021. Available genomes and plasmids were obtained from PATRIC and NCBI. Resistomes and methylomes were analysed using BAcWGSTdb and REBASE whilst Kaptive was used to predict capsule typing. Plasmids and other MEGs were identified using MGE Finder and ResFinder. Phylogenetic analyses were done using RAxML and annotated with MEGA 7.

A total of 877 isolates, 32 genomes and 44 plasmid sequences were analysed. Most of these isolates were reported in Asian countries and were isolated from clinical, animal, and environmental sources. Colistin resistance was mostly mediated by *mgrB* inactivation, while OXA-48/181 was the most reported carbapenemase. IncX and IncI were the most common plasmids hosting carbapenemases and *mcr* genes. The isolates were co-resistant to other antibiotics, with *floR* (chloramphenicol) and *fosA3* (fosfomycin) being common; *E. coli* ST156 and *K. pneumoniae* ST258 strains were common globally. Virulence genes and capsular KL-types were also detected. Type I, II, III and IV restriction modification systems were detected, comprising various MTases and restriction enzymes. The escalation of highly resistant isolates drains the economy due to untreatable bacterial infections, which leads to increasing global mortality rates and healthcare costs.

**Author summary:** Carbapenem and polymyxin co-resistance in Gram-negative bacteria (GNB) is a growing public health concern globally. China presented a high number of highly resistant bacteria from humans, animals, and environmental samples. *Escherichia coli* isolates were the most dominant in China, while *Klebsiella pneumoniae* isolates dominated Greece, the second country with high numbers of carbapenem and colistin co-resistant strains. Mobile genetic elements including plasmids are responsible for disseminating such resistance genes. Worryingly, GNB with Carbapenem and polymyxin co-resistance also harbours genes that make them resistant to other antibiotics, making them multi-drug resistant.

## 1. Introduction

Antibiotics resistance (AR) in Gram-negative bacteria (GNB) result in severe infections that are difficult to treat. Multiple AR genes (ARGs) in GNB, which confer resistance to two or more antibiotics, make the host bacteria multi-drug resistant (MDR) ^1, 2^. GNB are continuously evolving as new genetic elements emerges and outspreads ^3–5^, increasing the incidence and dissemination of MDR infections that pose a huge threat to public health globally^6^. High demands for and over prescription of antibiotics in humans and animals have resulted in an increasing burden of AR globally ^7, 8^. Increasing rates of AR in GNB have influenced and driven the need for novel antibiotics and newer treatment therapies ^9, 10^. Antibiotics such as cephems and carbapenems have successfully been used to treat infections caused by highly resistant GNB, particularly *Enterobacteriaceae* ^1, 11, 12^. However, enormous use of these antibiotics in human and veterinary medicine is resulting in the emergence of bacterial resistance to these important antibiotics ^13^. An even more concerning problem is that carbapenem-resistant GNB also develop resistance to other antibiotics, including polymyxins such as polymyxin E (colistin), which is regarded as the last “hope” and has been effective in treating infections caused by MDR GNB thus far ^14–16^.

Carbapenems are antibiotics that are reserved for treating infections caused by most infectious pathogens, including carbapenem-resistant *Enterobacteriaceae* (CRE^3, 13^. They are β-lactams with an extensive activity against infectious GNB, which cause complex, highly resistant infections ^1^. Carbapenems are hydrolyzed by enzymes known as carbapenemases that target the β-lactam ring of a β-lactam antibiotic to deactivate it ^17, 18^. Carbapenemases include the broad-spectrum New Delhi metallo-β-lactamase (NDM), Verona integrin-encoded metallo-β-lactamase (VIM), Imipenemase (IMP) ^19, 20^, Oxacillinases (OXA) ^21^, Guiana extended–spectrum β-lactamases (GES) and *Klebsiella pneumoniae* carbapenemase (KPC) ^17, 22^.

While there is still an ongoing global battle against acquired carbapenem resistance in GNB, polymyxin resistance is also emerging at a faster pace ^23^, owing to the global increase in the use of polymyxins to treat carbapenem-resistant infections ^14, 15^. Polymyxin resistance is primarily mediated by a mobile colistin resistance (*mcr*) gene ^14, 15^, as well as by modification(s) in or loss of lipopolysaccharides (LPS) and increase in capsule production, which often prevents polymyxins from binding to the target site ^23, 24^. Polymyxins are positively charged antimicrobial peptides and they target negatively charged LPS of the bacterial outer membranes ^25^. Notably, the *mcr* gene is mostly located on plasmids, giving them an advantage to spread to other bacteria easily and rapidly. Presently, ten variants of the *mcr* gene have been identified, with the *mcr-1* gene being the most common and first identified variant ^14, 15^ Chromosomally, polymyxin resistance occurs via active mutations in the two-regulatory systems, which in turn modifies the LPS ^24^. Several studies have now reported that mutations in the two-component systems; CrrA/B, PmrA/B and PhoP/Q and inactivation of the negative regulatory gene *mgrB* are connected to polymyxin resistance in GNB ^22, 24, 26–28^. These systems play a role in the modification of the LPS, which confers colistin resistance in GNB ^15^. The two-regulatory systems in GNB regulate cellular function based on various environmental signals including antibiotic exposure ^29^.

The *pmrAB* system is part of the *pmrABC* operon, which play a huge role in chromosomal colistin resistance. This operon encodes cytoplasmic membrane-bound sensor kinase (PmrB), regulatory protein (PmrA) and a putative membrane protein (PmrC) ^15^. LPS is negatively charged, which ideally increase binding affinity for positively charged colistin molecules. However, mutations on the two regulatory system modifies the LPS, reducing its overall negative charges. This in turn lowers the chances of colistin molecules to bind, making a bacterium resistant to colistin ^30^. Mutation(s) on *pmrA* and/or *B* genes may lead to the modification of Lipid A, which may results in colistin resistance ^29^. The *phoPQ* system regulates the *pbgP* operon^15^, which encode membrane enzymes that are responsible for the formation and addition Ara4N (4-amino-4deoxy-L-arabinose) to Lipid A ^31^. Such enzymes are EptA (phosphothanolamine) and ArnT (glycosyltransferase) and are responsible for the remodelling of the surface structure of the Lipid A by adding Ara4N to 4’ position of the Lipid A ^24^. Therefore, the expression of such gene’s triggers Lipid A modification, thus mediate bacterial polymyxin resistance. Furthermore, any changes on the *mgrB* and *crrB* regulatory genes also results in the activation of ArnT and EptA through the *arnBCADEF* operon respectively ^24, 25^.

Co-resistance to polymyxins and carbapenems is critical and more concerning as it threatens global antibiotic chemotherapy, patients’ recovery, and the economy ^3, 15^. Unfortunately, such bacteria also harbour other ARGs conferring resistance to aminoglycosides (*rmtA-H, armA, aac6’-IIb*), fosfomycin (*fosA*), cephalosporin (*bla_CTX-M_*), tetracycline (*tet*) and florfenicol (*floR*) ^16, 32, 33^. Hence, GNB with co-resistance to polymyxins and carbapenems are mostly always MDR. This crisis continues to challenge healthcare facilities, increasing morbidities, mortalities, and healthcare costs ^34^.

In addition to ARGs, bacteria have systems that allow them to survive antibiotic stress and other external aggression. Such systems include the production of protective capsules and restriction modification systems (RMS): DNA methylases (MTases) and restriction endonucleases (REs) ^35^. DNA methylation is the principal epigenetic modification in cellular processes including DNA replication and repair, lowering of transformation, transcriptional modulation, regulation of gene expression and defense against foreign DNA ^35^. DNA methylation is also important in regulating gene expression including that of resistance and virulence genes ^36^. MTases and Res have the same DNA recognition site (motif) and are frequently encoded alongside each other ^35^. Mtases methylates DNA by adding a methyl group and restriction endonucleases cleave exogenous DNA owing to different patterns of methylation ^37^. However, MTases can either act with or independent from restriction enzymes and such MTases are referred to as orphan MTases ^35^.

Although these processes are also found in eukaryotes, they are very diverse in prokaryotes and are categorized as Type I, II, III and IV ^35, 37, 38^. These classifications are differentiated by recognition site, cleavage site, subunit composition and the type of substrate ^38^. Type I and II are the most prevalent systems in bacteria. The Type I system constitute three subunits, which are responsible for specificity (S), modification (M) and restriction (R) ^35, 38^. On the other hand, the Type II system has the R and M subunits only ^38^. The R and M genes in type II system are separate but has the same binding site ^38^. The type II restriction enzymes recognize and destroy acquired and/or foreign DNA with either incomplete or absence of methylation signature (motif) ^39^.

Mtases may be associated with mobile genetic elements (MGEs) such as plasmids and transposons that contribute to the difference in bacterial methylation patterns ^35^. Furthermore, the RMS may also be encoded by genes found on chromosome or plasmids, which are also responsible for disseminating ARGs ^40^. Hence, plasmids without the same methylation signatures as the host cell are regarded as “foreign” and will be susceptible to cleavage by restriction endonucleases ^39^.

Bacteria also produce and secrete extracellular material called polysaccharide capsules which allow them to bypass the host immune system ^41^. These are usually made of a chain of simple sugars attached to the outer surface of bacterial cells and are therefore the first elements to interact with the host ^41^. The formation of these capsules is linked to efflux pumps, which are pervasive membrane transporter proteins and are important for bacterial antibiotic resistance ^42–44^. These systems pump out toxic substances/solutes such as metabolites and antimicrobial agents from the cell ^42^. Efflux pump such as knpEF, a regulatory member of small multidrug resistance protein (SMR) group mediates polymyxin resistance ^42, 43^. This pump is responsible for the formation of capsular polysaccharides in *K. pneumoniae* ^42–44^. When this pump is activated, more capsules are synthesized leading to an increase in colistin resistance as they serve as a barrier between colistin and bacterial cells ^42^. Therefore, resistance level in GNB may depend on the number of capsular polysaccharides produced ^42^. Furthermore, tighter encapsulation in *K. pneumoniae* also contribute to the increase in polymyxin resistance. This occur through the inactivation of the *mgrB* gene, resulting in harder and tightly bound capsular polysaccharides ^45^.

Antibiotic resistance is commonly disseminated through MGEs including transposons, plasmids, and insertion sequences ^40^. Plasmids are the most common vehicle for resistance genes transmission, offering an advantage to bacteria as it may bypass the species barrier^3, 4, 15^. Therefore, plasmid borne ARGs such as *mcr* and carbapenemases are widespread due to their faster dissemination rate among bacteria species ^40^. Owing to the close connection between humans, animals, and the environment (water, soil, air, and communities/built environment), ARGS and virulence mechanisms can be disseminated between these three domains through bacterial hosts and plasmids ^7, 8, 46^. Thus, antibiotics administration in livestock/food-producing animals may select and spread antibiotic-resistant GNB to humans and the environment through the food chain ^7, 8, 46^ and their faeces/farm effluents, respectively. Therefore, applying the One Health concept, which describes the interaction between humans, animals, and the environment, may be effective in reducing the spread of ARGs between these domains ^47^. Herein, we present an epidemiological and molecular analysis of GNB that are co-resistant to carbapenems and polymyxins from a One Health perspective.

### Evidence before this review

Dandachi et al. (2018) published a review based on a study done in the Mediterranean basin, which only focused on co-resistance of carbapenem and colistin among GNB from animal samples ^48^. Thus, the study was only based on a specific region and samples were isolated from one source (animals) only. Furthermore, the study did not address the epigenomics, genomics, virulome, and evolutionary phylogenomics of the resistant isolates. So far, not much is known about the depth of the genetic structure of GNB with carbapenem and colistin co-resistance; such evidence is still missing. Another review article on a multidrug-resistant GNB done in the Middle-East was published ^49^. Although this study reported from a One Heath view from a region, it did not look at the epigenomics and evolutionary phylogenetics of the isolates.

### Purpose of this systematic review

This systematic review and genomic analyses report on a One Health epidemiology of colistin resistance in carbapenemase-producing GNB on a global scale. Further, the epigenomics, mobilome, capsules and virulome of GNB genomes with carbapenem and colistin resistance mechanisms are described alongside their geographic location, sample source, and other ARGs.

## 2. Methods

### 2.1 Literature search

An extensive literature search was done on PubMed using the following search words/phrases: carbapenem* AND colistin/polymyxin AND resistan* AND Gram-negative bacteria; carbapenem* AND colistin/polymyxin AND resistan* AND bacteria; and carbapenem* AND mcr AND resistan* AND bacteria. The search was done in a factorial fashion. Other search engines used included google scholar, ScienceDirect and ResearchGate, which resulted in twelve additional papers.

#### 2.1.1 Inclusion and exclusion criteria

Articles published between January 2016 and March 2021 were included. A total of 1769 articles that did not address GNB and were excluded (Table S1_A). Further screening excluded 66 articles, which focused on the resistance to one antibiotic. A total of 129 articles, which focused on carbapenem and polymyxin co-resistance in GNB, were included for qualitative synthesis and statistical analyses (Table S1_B).

### 2.2 Collection and characterization of isolates in included studies

For studies included in this review, samples were broadly collected from hospitals, farms, and animal slaughterhouses. GNB were isolated from bodily fluids, wounds, catheter tip, floor, sewage as well as from food. After sample collection, bacteria were cultured, followed by DNA extraction, gene amplification and whole-genome sequencing. For antibiotic susceptibility tests (AST), most studies employed the broth microdilution method (BMD) to determine colistin resistance ^19, 22, 50–52^. The genomic sequences were also used to identify carbapenemases and colistin resistance mechanisms, as well as assign sequence types (ST) of clones based on multi-locus sequence typing (MLST) ^53^. Conjugation assays were used to assess the transferability of carbapenem and colistin ARGs followed by the selection of transconjugants on various agar plates containing various antibiotics including carbapenem and colistin.

For this study, bioinformatics and statistical analyses of downloaded genomes and extracted data were undertaken, respectively. Information such as the geographical distribution, sample source, species, clones, ARGs, MGEs and plasmids replicons/incompatibility (Inc) groups was extracted. A total of 877 GNB isolates from different sources were analyzed (Table S2_A).

### 2.3 Statistical analysis

Data from 129 articles was analyzed using Microsoft Excel 365. Differences and patterns across different data variables such as countries, species, specimen sources, ARGs, MGEs etc. were also analyzed using Microsoft Excel 365. Furthermore, results were visualized through bar charts, pie charts and maps from Microsoft Excel. Statistical significance between different data sets and variance analysis were calculated using One-way ANOVA and/or One Sample T-test on GraphPad. A probability value (p-value) of <0.05 was defined as a significant correlation between different datasets.

### 2.4 Included Genomes

Whole genome (chromosome and plasmid) sequences of isolates with both carbapenem and colistin resistance were downloaded from the NCBI (https://www.ncbi.nlm.nih.gov/) and PATRIC (https://patricbrc.org/) bacterial genome databases, using the accession numbers provided in the included articles. A total of 32 whole genome sequences and 44 plasmids were retrieved. Of the 44 plasmids, 28 and 16 were respectively associated with carbapenemases and *mcr* genes, respectively. Other isolates were not included in the analysis because their accession numbers were either not provided or not found on the databases.

### 2.5 In silico analyses of included genomes

For the available genomes, the DNA methylases (methylome), motifs, RMS were identified on the restriction enzyme databases (REBASE) (http://rebase.neb.com/rebase/rebase.html) hosted at the Centre for Genomic Epidemiology. ARGs and plasmids replicon were determined using ResFinder (https://cge.cbs.dtu.dk/services/ResFinder/) and plasmidFinder (https://cge.cbs.dtu.dk/services/PlasmidFinder/) whilst the virulomes were identified using BAcWGSTdb (http://bacdb.org/BacWGSTdb/). Other MGEs were identified using using MGE Finder (https://cge.cbs.dtu.dk/services/MobileElementFinder/). The evolutionary phylogenetics of the isolates were analysed on RaxML (https://raxml-ng.vital-it.ch/<u>#/</u>). Phylogenetic trees were visualized and annotated using Molecular Evolutionary Genetics Analysis version (MEGA) 7.0 ^54^. Predictions of capsules polysaccharides were done on the Kaptive WEB (https://kaptive-web.erc.monash.edu/) based on the K-type serotyping of *K. pneumoniae* ^55^.

## 3. Results

### 3.1 Global distribution of colistin and carbapenem co-resistant GNB isolates

Literature search resulted in a total of 3048 articles published in English with two duplicates, resulting in 3046 articles for screening. Further review and sorting resulted in 195 articles of which 66 were excluded and the remaining 129 articles were used or meta-analyses. (Figure 1). A total of 877 isolates from 43 countries were included for analyses. These constituted various species including *Klebsiella* spp. (n = 536), *Escherichia coli* (n = 210), *Acinetobacter* spp. (n = 71), *Enterobacter* species (n = 20), *Pseudomonas aeruginosa* (n = 18), *A. calcoaceticus-A.baumannii* complex (n = 13), *Cronobacter sakazakii* (n = 2) and *Citrobacter amalonaticus* (n = 1) (Figure 2A).

**Figure 1:**
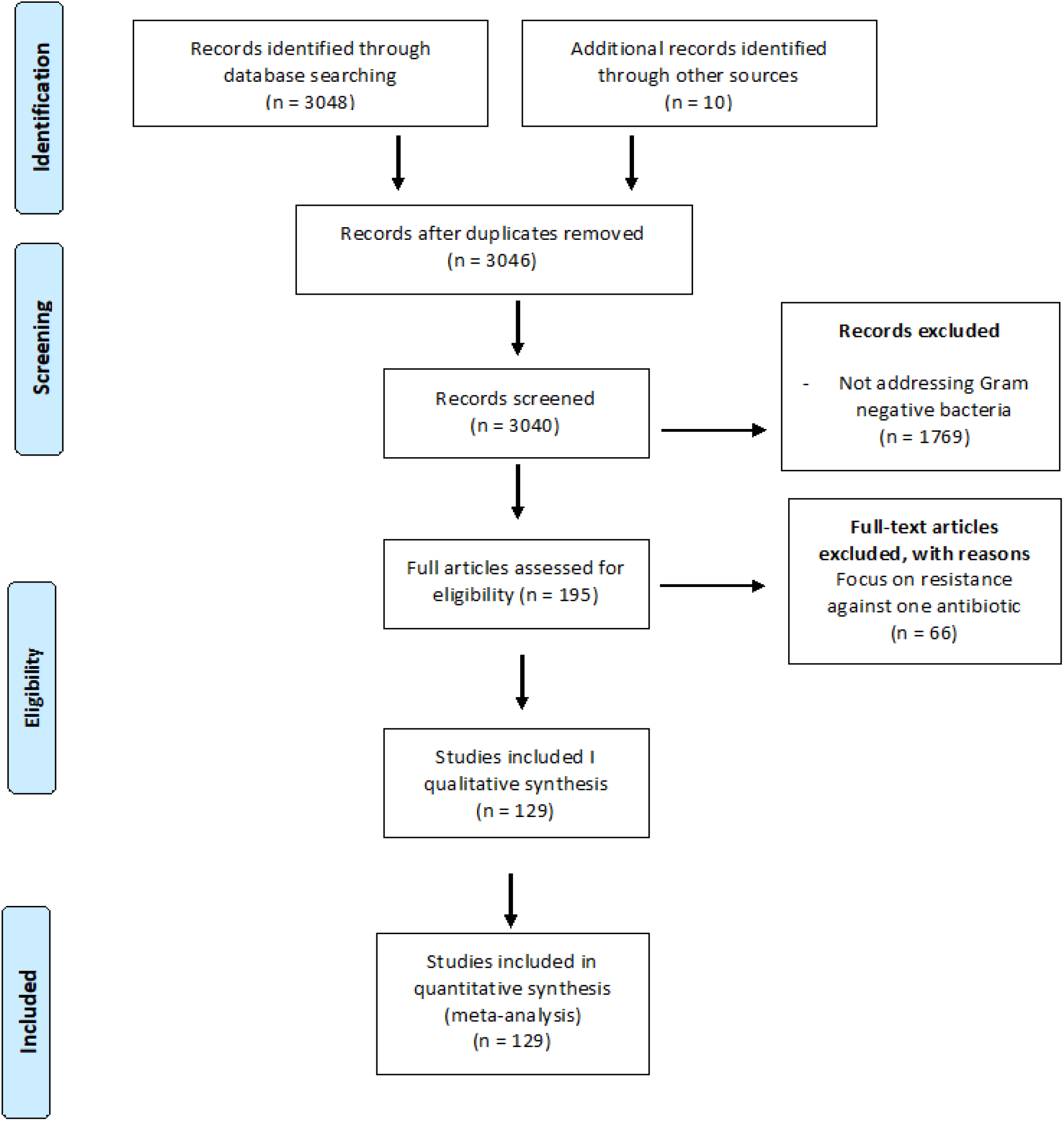
A flow chart showing the literature search strategy, inclusion, and exclusion criteria used for sorting the data.

**Figure 2:**
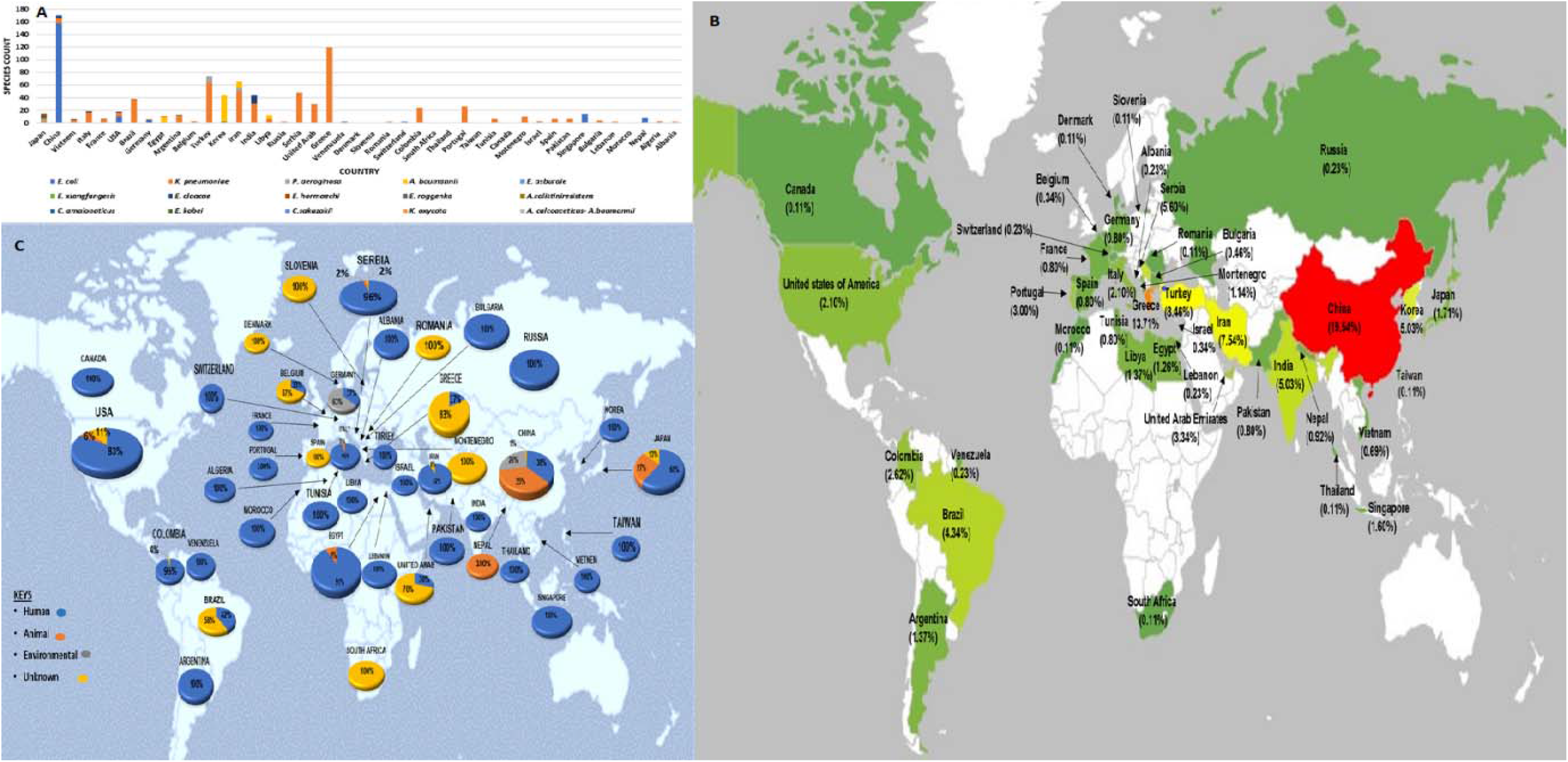
Global distribution of species and sample sources from which carbapenem and polymyxin-resistant Gram-negative bacteria were isolated from. (**A**) shows the global species distribution of GNB with carbapenem and colistin co-resistance per country, (**B**) Global distribution of carbapenem and colistin co-resistance in GNB. The scale ranges from a country with the highest prevalence (red) to the one with the lowest distribution (green) and (**C**) shows species count in different countries based on the source of isolation.

Carbapenem and polymyxin co-resistance in GNB was detected in five continents across the globe, except for Australia (Figure 2B). Asia presented high numbers of carbapenem and polymyxin co-resistance (n = 415), followed by Europe (n = 333), South America (n = 76), Africa (n = 35), and North America (n = 9) (Figure 2B). As shown in Figure 2B, cases of emerging MDR GNB with co-resistance to carbapenem and colistin have mostly been reported in Asian countries. China, having the highest cases globally and in Asia, reported 19.54% MDR GNB isolates (n = 171/877) of MDR GNB with carbapenem and polymyxin co-resistance. Although co-resistance to last-resort antibiotics in GNB is more prevalent in China, it is evident that this trend has spread beyond Asian countries into European countries such as Greece and Portugal representing 13.7% (n = 120) and 3.0% (n = 26), respectively.

Asia, Europe, and North America reported carbapenem-polymyxin resistance in GNB from animals while Africa and South America only presented GNB isolates from humans. Overall, human samples globally constitute 65.5% (n = 574) while animals, environment and unknown sources represents 8.7% (n =76), 5.7% (n =50) and 20.2% (n = 177), respectively (Figure 2C). The statistical analysis of all sample sources was significantly reported in different countries (p = 0.02) (Table S2_B).

China presented the most diverse specimen sources than other countries, with four species. China reported resistant isolates from all samples sources but showed a non-significant association among sample source (p = 0.75) Table S2_C). Figure 2C shows that of the cases reported in China, 37.4% (n = 64) of the isolates were from human samples, 35.08% (n = 60) from animals, two from unknown sources (1.2%), and 26.3% (n = 45) from environmental sources including slaughterhouses, farm, water, sewage etc. Furthermore, Liu B et al (2017) and Wang R et al (2018) reported a total of 14 and 26 isolates from animals’ source, respectively ^6, 56^. Of the isolates reported by Wang R et al (2018), three isolates (*E. coli* WFA16004, *E. coli* WFA16019 and *E. coli* WFA16061) presented a new sequence type. A total of 18.0% (n = 21) of the cases in Greece were from human samples, and the remaining 0% (n = 99) were of unknown sources. Iran (7.52%., n = 66) and Korea (5.01%, n = 44) also presented more cases of carbapenem and colistin co-resistance in Asia^6^. In contrast to the cases reported in China, Iran and Korea do not have any animal or environmental cases of GNB harbouring carbapenem and polymyxin ARGs (Figure 2C).

### 3.2 Resistomes of GNB with carbapenem & polymyxin co-resistance

#### 3.2.1 Carbapenem & polymyxin resistance mechanisms

From a total of 877 reported GNB isolates, there was a significant association between ARGs/resistance mechanisms in different countries (p = 0) Table S2_D). Colistin resistance through the inactivation of the *mgrB* gene presented 37.3% (n = 322), while resistance through *mcr-1* gene presented 36.2% (n = 312) (Figure 3A). Resistance via mutation of the regulatory systems was 5.10% (n = 44) and 3.82% (n = 33) for *prmA/B/C* and *phoP/Q,* respectively. Furthermore, six isolates were resistant to colistin via mutation on the *crrB* gene: one in Colombia (*K. pneumoniae* C7), France (*K. pneumoniae* 20.70), Greece (*K. pneumoniae* G104), and South Africa (*K. pneumoniae* Af44b) and two from Brazil (*K. pneumoniae* CrB8i & CrB9i) (Table S2_A). A total of 146 isolates (17.0%) were resistant to colistin, but their resistance mechanisms were not specified. Of the undetected mechanism, one strain, *A. colistiniresistens* from a human sputum in Japan, was reported to be naturally resistant to colistin ^57^. *mcr-1*-mediated colistin resistance was more prevalent in *E. coli* isolates (p = 0.72) while resistance via mutations in the regulatory systems was only reported in *K. pneumoniae* (p = 1.00) and *A. baumannii* (p = 1.00) Table S2_E). *mcr-1* was frequently associated with IncX_4_ replicon plasmids in *E. coli* species ^58, 59^. Another variant in the *mcr* family, *mcr-9,* was detected in *K. pneumoniae* isolates and was more prevalent in European countries including Montenegro (n = 10) and Spain (n=7) ^20^. This gene was also reported in Japan (n = 3) ^57^ (Table S2_A). Other identified *mcr* variants included *mcr-1.2* reported in Italy ^33^, *mcr-1.5* ^60^ from Argentina and *mcr-3* ^60^, *3.5,* and *4.3* ^61^, and *mcr-8* ^62^ from China. These were each detected from human samples; except *mcr-8* which was isolated from a swine in China (Table S2_A).

**Figure 3:**
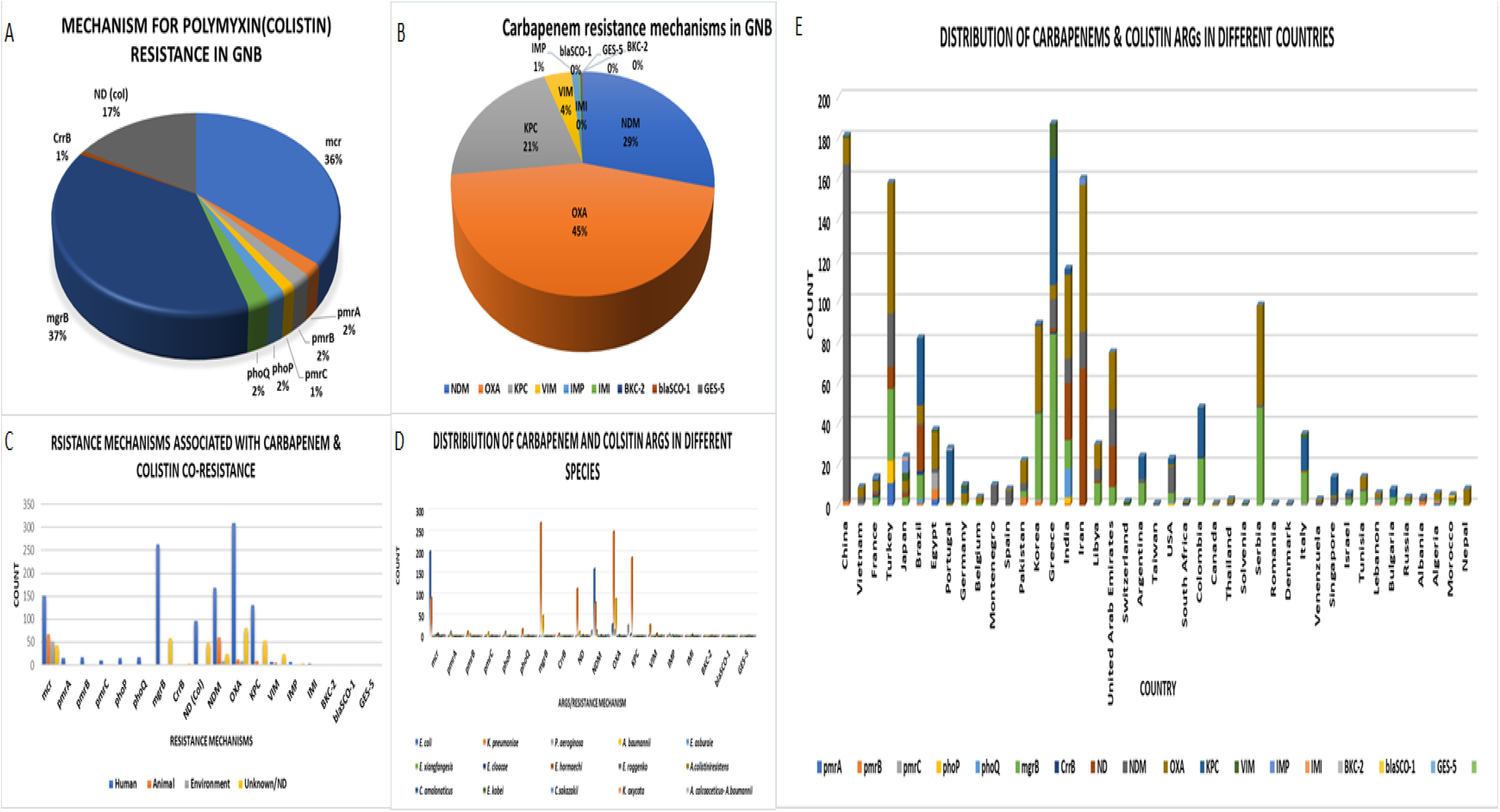
Types and distribution of colistin and carbapenem resistance mechanisms and associated resistance genes. **(A)** Resistome dynamics of colistin in GNB that are co-resistant to carbapenem and colistin. (**B**) Carbapenem resistance mechanisms in GNB that are co-resistant to carbapenem and colistin. (**C**) shows resistance mechanisms based on species source. (**D**) shows carbapenem and colistin resistance mechanisms in different species. (**E**) shows the distribution of carbapenamases and colistin ARGs in different countries.

Carbapenemases such as NDM, OXA-48 type, IMP, VIM, GES, SCO, BKC and KPC were reported, where OXA-48 type, NDM and KPC were the most common carbapenemases (Figure 3B). The most common carbapenemases reported globally was OXA-48 type presenting 45.0% (n = 411) of 877 isolates and this was prevalent in *K. pneumoniae* isolates (n = 249). This was followed by NDM 29.0% (n =261), KPC 21.0% (n = 192), and VIM 4.0% (n = 37), IMP 1.0% (n = 10). IMI (n = 3), GES-5 (n = 1), SCO-1 (n = 1) and BKC-2 (n = 1) were less the 1% of carbapenemases (Figure 3B). The most common variant of NDM was *bla*_NDM-5,_ which was more prevalent in *E. coli* isolates from China. OXA-48 was mostly reported in *K. pneumoniae* and *P. aeruginosa* isolates, while OXA-23 was found in *A. baumannii* (in Seoul, South Korea) and in *A. calcoaceticus-A. baumannii* complex (in India) from human samples (Table S2_A). Notably, *mcr-1* mostly co-occured with the *ndm-5* and/or *ndm-9,* mostly in *E. coli* isolates. Furthermore, colistin resistance by mutation co-existed wit *kpc* in *K. pneumoniae* isolates.

#### 3.2.2 ARGs associated with polymyxins and carbapenem co-resistance in GNB

The resistance profiles of the isolates in Figure 4A & B indicated resistance to a broad spectrum of antibiotics including penicillins, cephalosporins, aminoglycosides, fluoroquinolones, tetracyclines, sulphonamides, and rifampins ^16, 22, 32, 63^ The expression of these genes would be problematic as it increases MDR and extensively drug-resistant (XDR) GNB, thus threatening global health.

**Figure 4:**
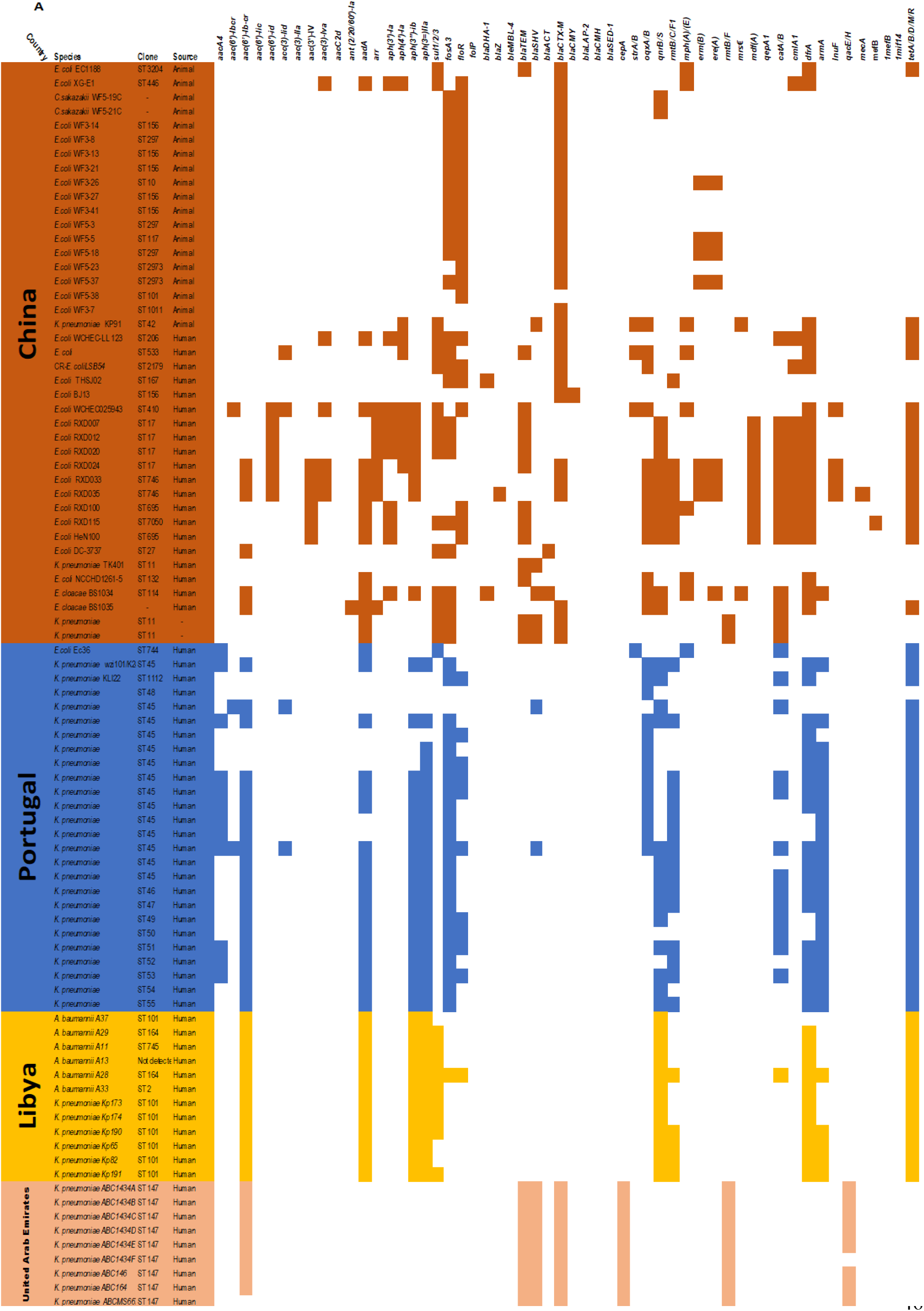
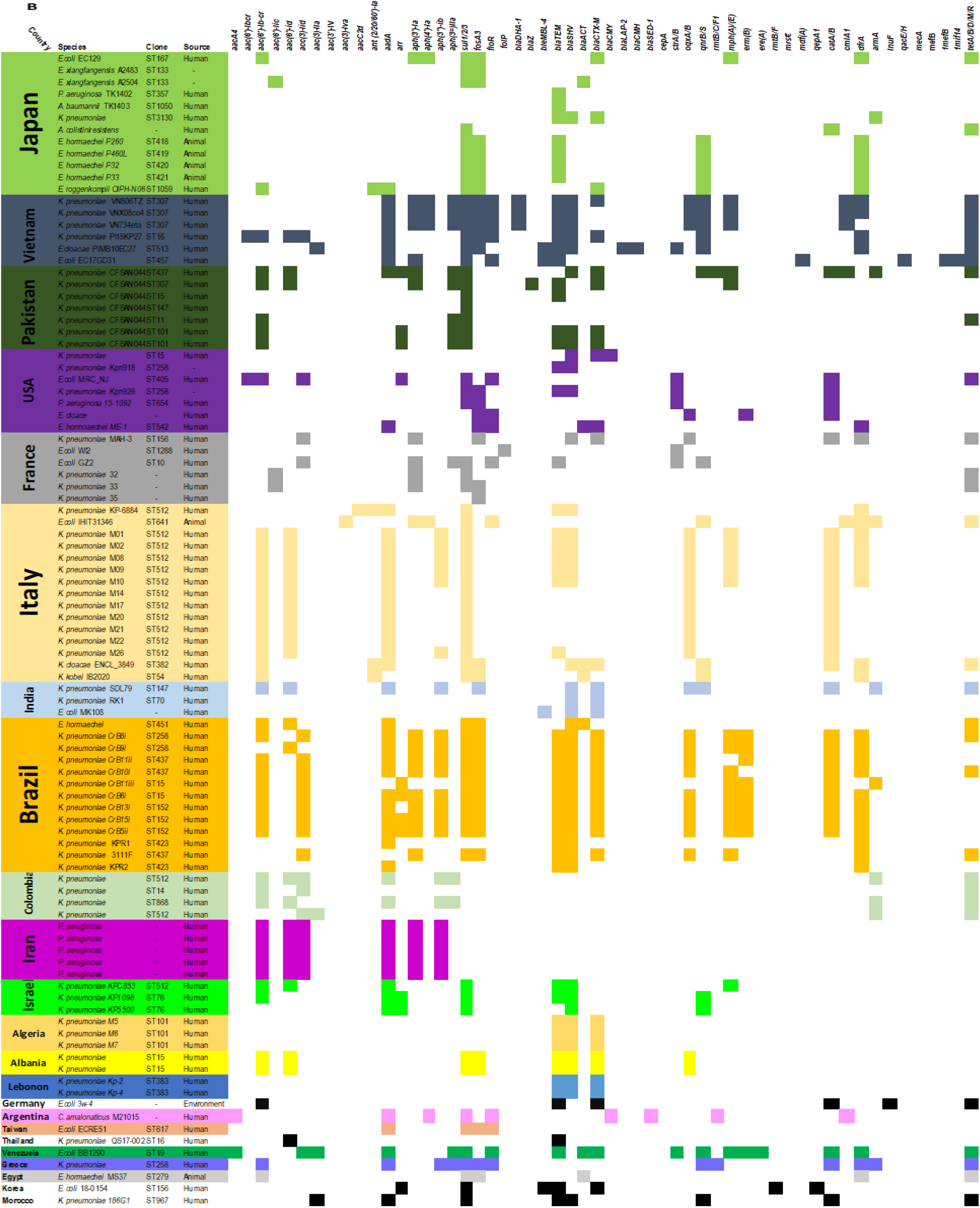
Global genomic resistome analysis of Gram-negative bacteria with carbapenem and polymyxin co-resistance genes. (**A**) and (**B**) both show the global distribution of other ARGs in addition to carbapenamases and colistin resistance genes in GNB in different countries.

Resistance profiles of 185 isolates are presented in Figure 4A & B. Other isolates were not included because of the unavailability of genome accession numbers and lack of such information from articles. The overall analysis of the resistance profiles showed that *sul1-3*, *floR* and *fosA3* genes, which codes for sulfamethoxazole, chloramphenicol and fosfomycin resistance respectively, were commonly associated with carbapenem and polymyxins resistance determinants. This was particularly true for most *E. coli* isolates globally, whilst extended-spectrum β-lactamases (ESBLs), viz. *bla*_SHV_ and *bla*_CTX-M_, were more common in *K. pneumoniae* globally, except for *K. pneumoniae* isolates in Portugal. The abundance of other antibiotics’ ARGs associated with those of carbapenem and colistin were higher in China, Pakistan, and Vietnam, suggesting that MDR genotypes are rapidly increasing and spreading in Asian countries than on other continents.

As shown in Figure 4A & B, *bla*_CTX-M_ was found in E. *coli* isolates from animals in China, but it was absent in *E. coli* IHIT31346 from animal sample in Italy. This gene was also detected in *E. coli* isolates from human samples from China (n = 6) and single isolates from France, Venezuela, Korea, Russia, and Japan. It was also found in *K. pneumoniae* from 10 different countries, and in *E. coli* 3w-4 from an environmental sample from Germany (Figure 4 A & B). Other common ARGs found in the 19 *E. coli* isolates from animals include *floR* and *fosA3*. Furthermore, three of the 19 *E. coli* isolates from animals also harboured the *erm(B)* gene, which confers resistance to macrolides. Notably, isolates from animals, particularly in China, do not harbour many ARGs alongside carbapenemases and polymyxin ARGs. Except MDR *K. pneumoniae* KP91, only one isolate from animal origin in China, carried more diverse ARGs including *bla*_SHV,_ *tet*, *strA/B*, *qxA/B*, *rmsE*, *mph(A)/(B)*, *sul1-3*, and *aph(4’)-Ia*.

On the contrary, *E. coli* isolates from humans in China showed more diverse distribution of ARGs (Figure 4A & B). However, this was not the case for *E. coli* BJ13 and NCCHD1261-5, which only harboured two β-lactamases; same was observed for *K. pneumoniae* TK401. As observed in China, human isolates from other countries also showed diverse ARG distribution than isolates from animals and the environment. Nonetheless, one environmental isolate from Germany, *E. coli* 3w-4, harboured more ARGs including *tet (B)*, *aac(6’)-lb-cr, catB*2, & *catB, drfA17,* and *bla*_CTX-M_ (Figure 4B). As shown for *E. coli* isolates in China, the *K. pneumoniae* isolates from human samples in Pakistan and Vietnam were resistant to a broad spectrum of antibiotics including quinolones (*aac(6’)-lb-cr*, *oqxA*/*B*, *qnrB/S*), tetracycline (*tet*), and aminoglycoside (*aac(3)-lla*, *aac(6’)-lb-cr*, *aadA*, *aph(3’)-lb*, *aph(6)-ld*) (Figure 4A & B). An uncommon resistance gene, *cmIA5,* which is responsible for chloramphenicol resistance, was detected in *K. pneumoniae* isolates (n = 3) from human samples (Vietnam) and in *E. coli* IHIT31346 of animal origin (Italy) ^64^. Furthermore, this gene was also reported in abundance in China (n = 12) from E. *coli* isolates from animal, human, and environmental samples, implying the presence of chloramphenicol resistance in Asian and European countries. In China, human isolates generally had a rich resistome repertoire than isolates from animals and the environment.

Libya reported a total of 12 isolates from human samples, all of which harboured the following genes: *tet*, *aadA*, *aac(6’)-lb-cr*, *aph(3=)lla, aph(3=)-lb,* and *qnrB/S* (Figure 4A). An isolate reported in Libya, *A. baumannii* 28 (ST164) was highly resistant, carrying additional ARGs including those against phenicol (*floR, catA/B*) and sulphonamides (*sul1*-3). Furthermore, human isolates in Libya and Portugal lacked ESBL genes except *K. pneumoniae* ST45 in Portugal from which the *bla*_SHV_ gene was found. Unlike isolates in Libya, isolates from Portugal did not harbour *sul1*-3 genes. However, the *sul1*-3 genes and additional ARGs such as *strA/B* and *mph(A)/(E)* were detected in *E. coli* Ec36 from a human sample in Portugal (Figure 4B). Further analysis of the resistance profiles revealed that multidrug resistance genotypes are generally not common in the US as the reported isolates only harboured ESBLs genes. This might be due to the number of carbapenem and colistin co-resistant isolates reported in the country, which calls for further studies.

### 3.3 Mobilome

#### 3.3.1 Plasmid profiles of GNB with carbapenem and colistin resistance

(Table S3_D) shows a total of 18 plasmid incompatibility groups. Plasmid replicons associated with carbapenem and colistin resistance genes reported globally includes IncI2/3, IncX3/4, IncH12, IncF/IIK/II/IB, IncN, IncA/C, IncL/M, IncP, IncR, IncY, IncA/C2, IncR, IncY and IncB/O (P= 0.002) (Table S3_B). Additionally, some countries reported hybrid plasmids, which harbors ARGs for different antibiotics. So far, China is the only country reporting the co-existence of carbapenem and colistin ARGs on a hybrid plasmid ^65^. The *bla*_NDM-5_ and *mcr-1* genes were both carried by a single replicon hybrid plasmid, IncX_3_-X_4_ identified in *E. coli* CQ02-121 which was isolated from a patient’s rectal swab from a health facility in China ^65^.

The IncX plasmids are widely reported as the most common MGE of carbapenems and polymyxins ^66^ (Figure 5A). Various groups of IncX plasmids, including IncX_1_-IncX_8_ have been identified as carriers for transmissible carbapenem and polymyxin ARGs ^12, 67^. The most prevalent IncX plasmids associated with *bla*_NDM_ (p = 0.107) and *mcr-1* (p = 0.999) ARGs (Table S3_B). IncX_3_ and IncX_4_, responsible for the transmission of *bla*_NDM_ and *mcr-1* genes respectively ^16, 64, 65^. The most common replicons for all isolates were IncX plasmids (n = 107) comprising IncX_4_ (n = 57), IncX_3_ (n = 49) and IncX_1_-N (n = 1). This was followed by IncI (n = 58), IncF (n = 50), IncH (n = 45), and IncB/O (n = 14) (Table S3_D). The least reported replicons were IncP, IncL, IncR, and IncF_1_, IncI_1_ and IncN, which were each reported in one or two isolates. The most common replicons, IncX_3_ and IncF, were associated with carbapenemases, while IncX_4,_ IncI and IncH were mostly associated with *mcr-1*.

**Figure 5:**
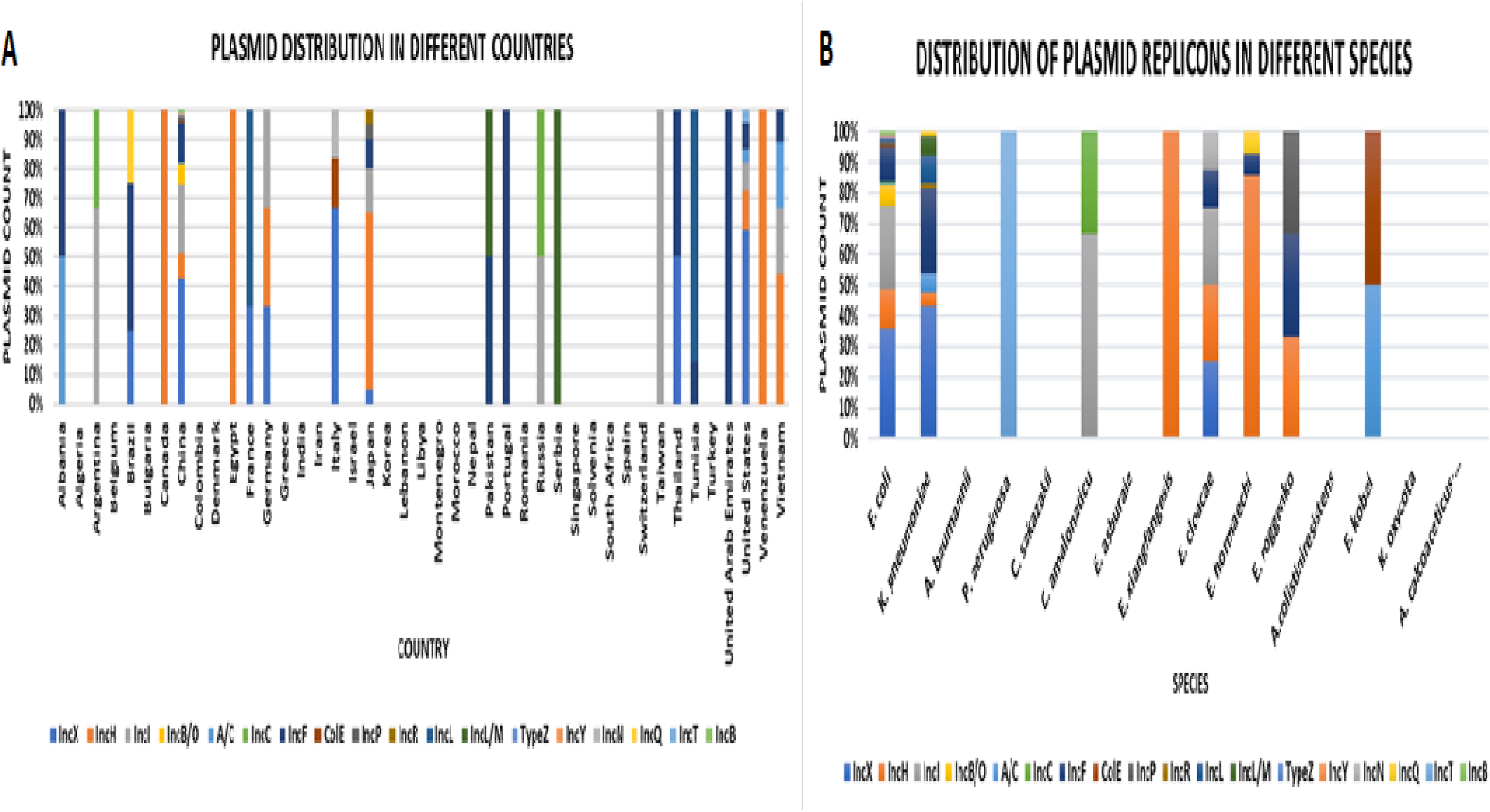
Global distribution of plasmid types found in Gram-negative bacteria with carbapenem and polymyxin co-resistance genes. (**A**) shows plasmid replicons associated with carbapenemases and colistin ARGs in different countries. (**B**) shows plasmid replicons in different species.

As expected, China reported a diverse plasmid replicon due to the high number of reported isolates with carbapenem and polymyxin co-resistance (n = 171). Therefore, IncX plasmid groups were commonly found as the carrier of resistance genes in China (Figure 5A): China reported a total of 84 IncX plasmids which were mostly reported in *E. coli* isolates (n = 72) (p = 0.138) (Table S3_B). Although *K. pneumoniae* in China did not harbour IncX plasmids, 25 *K. pneumoniae* human isolates in Portugal had their *mcr-1* gene carried by IncX_4_ plasmids (Figure 5B). The second most common plasmid was IncI, especially IncI_2,_ which carried *mcr-1*and was commonly reported in China (n = 39).

IncL/M was only reported in Pakistan (n = 5) and it carried *bla_OXA_* genes in *K. pneumoniae* isolates (p = 0.057), while IncB/O replicon was only reported in *E. coli* (p = 0.138) isolates from animal samples in China (Table S3_B). IncF carried *bla_NDM_* in various countries, whilst IncR was associated with various carbapenemases such as *bla_NDM-1_* in *E. coli* isolates from Vietnam (n = 1) and China (n = 1). It was also found with *bla_KPC-2_* in *E. coli* H17 from USA and *bla_OXA-181_* in *E. coli* IHIT31346 from animal sample in Italy. Another least common plasmid replicon associated with *bla_NDM_* in *E. coli* isolates was IncP, reported only in China and Japan (Figure 5B).

#### 3.3.2 Other MGEs associated with carbapenem and colistin resistance

In addition to plasmids, other MGEs associated with carbapenem and polymyxin resistance in GNB were reported. Such MGEs were 135 insertion sequences (IS), 60 transposons (Tnp) and four class I integrons (Table S3_A). The most prevalent IS was IS*903* (n = 28) and the most prevalent Tnp was *tnp*4401d (n = 31). Both IS*903* (n = 108) and *tnp*4401d (n = 74) were more common in *K. pneumonia* isolates (Table S3_A). The IS*903* was found within the *mgrB* gene in two *K. pneumoniae* isolates from the United states ^27^. *mcr-9* was flanked by IS*903*, IS1 or cupin fold metalloprotein (WbuC) in ten *K. pneumoniae* isolates in Montenegro; as well as in Serbia (n = 1) and Belgium (n = 1). Likewise, these MGEs were further associated with *mcr-9* in *K. pneumoniae* isolates in Spain (n = 7), Slovenia (n = 1), and Denmark (n = 1) (Table S2_A). *Tn4401d* was reported in *K. pneumoniae* isolates in Portugal (n = 21), USA (n = 2) and China (n = 1). This MGE was associated with *bla_KPC-3_* in Portugal, *mcr-8,* and *bla_NDM_* in China; and *bla_KPC-2_* in the United States (Table S2_A). Three class I integrons were reported in two *K. pneumoniae* isolates with *bla_KPC-2,_* in Brazil and in *A. colinistiniresistens* from Japan, which is naturally resistant to colistin ^57^ and harboured *bla_IMP-34_* and *bla_OXA-58_.* In416 integron was identified from *E. hormaechei ME-1* isolate carrying VIM-4 in USA (Table S2_A).

*ISAba*125, was associated with *bla_NDM-5_* in seven *E. coli* isolates from animal samples in China. This IS was also found in *Enterobacter cloacae* PIMB10EC27 from a human sample in Vietnam, flanking *bla_NDM-1_*; *E. cloacae* PIMB10EC27 further harboured *ISIR*21, *ISKpn*19 and Tn*6901* ^68^. *IS*5 was reported in three *Enterobacter* strains from Japan where it was associated with *bla_IMP-1,_* as well as in two *K. pneumoniae* isolates where it was associated with *bla_KPC-2_* and *bla_NDM/OXA_,* from Brazil and Pakistan, respectively. Other reported IS includes *ISApI*1 in *A. baumannii* TK1403, which existed alongside *bla_OXA_* genes, ISR in *K. pneumoniae* KP7 occurring with *bla_KPC_* (Table S2_A).

*ISAba*125 is the main carrier of *bla*_NDM_ ^69^, evinced by its appearance in two *E. coli* isolates facilitating the dissemination of *bla*_NDM-5_ and *bla*_NDM-6_ genes ^70^. Ho et al (2018) also reported on *IS*26-*I*Aba125 transposon complex that is associated with *bla*_NDM-5_ in isolates from animal origin^71^. *E. coli* Ec36 isolated from human urine in Portugal harboured *IS*Kpn7, *IS*Kpn6 and TN*40* ^72^. The most prevalent MGE on plasmids harbouring *mcr-1* was the insertion sequence *IS*ApI1 reported in China, Vietnam, Brazil, and Argentina (Table S2_A). The *IS*ApI1 insertion is normally responsible for mobilising *mcr-1* in GNB ^40^. In China, this MGE was associated with *mcr-1* from *E. coli* HeN001 and *C. sakazaki* WF5-19C (Table S2_A). It was also associated with the acquisition and dissemination of ARGs from animal samples and was identified in *K. pneumoniae* isolates in Brazil and Vietnam from human samples. Further, it was associated with the IncI_2_ replicon, which carried both *mcr-1.5* and *bla*_NDM-1_ in *C. amalonaticus* isolates from human samples in Argentina.

### 3.4 Capsular polysaccharides and virulome of GNB isolates

Table S4 shows various plasmid-mediated and chromosomal-borne virulence genes in MDR GNB. Virulence genes were identified in 15 *K. pneumoniae*, 12 *E. coli* and one *E. hormaechei* isolates resulting in a total of 29 genes (Table S4). The most common virulence gene in *E. coli* isolates was *gad,* a chromosomal glutamate decarboxylase found on the outer membrane. *E. coli* EC17GD31, isolated from human blood in Vietnam harboured 18 chromosomal virulence genes, except for outer membrane protein complement resistance (*TraT*) (Table S4). These isolates also had two copies of the *gad* gene. *E. coli* MCR-1_NJ, isolated in the United States, also harboured 18 virulence genes with duplicates of *Trat*, *terC* (tellurium ion resistance protein) and *KpsE* (capsule polysaccharide export inner-membrane protein) (Table S4). *E. coli* WI1 had a unique virulome structure, possessing unique virulence genes such as colicin B (*cba*), colicin ia (*cia*) and colicin M (*cma*), which were only shared with *E. coli* WI2 (Table S4).

For *K. pneumoniae* isolates, ferric aerobactin receptor (*iutA*) was commonly recorded *K. pneumoniae* KP-6884 and QS17-0029 harboured high numbers of virulence genes (n = 6). The least number of virulence genes were observed in *K. pneumoniae* CFSAN04457, 65, 73 (n = 3) and *E. hormaechei* ME-1 (n = 1). All virulence factors in *K. pneumoniae* QS17-0029 were found on the plasmid while KP-6884 had two *iutA* on the chromosome (Table S4). *K. pneumoniae* KP-6884 also presented a case where the same gene appeared both on a chromosome and plasmid (Table S4). Likewise, *traT* in *K. pneumoniae* isolates was also found on the plasmid. However, this gene was missing in *K. pneumoniae* CFSAN044572, CFSAN044573 and CFSAN044563 isolates.

Capsular polysaccharides were determined for 12 *K. pneumoniae* and one *A. colistiniresistens* isolates (Figure S1). The *Klebsiella* K-locus primary reference and the *Acinetobacter* K-locus primary reference were used to predict capsules for *K. pneumoniae* and *A. colistiniresistens* isolates, respectively. Eight KL-types and zero O-types were identified for *K. pneumoniae* isolates. The KL serotypes KL103, KL51, KL17 and KL2 were each identified in two isolates while KL112, KL107, KL102 and KL24 were present in one isolate, each (Figure S1). The capsular loci had between 15 and 23 genes, with two allelic types: *wzc* and *wzi.* The allelic types were present in all isolates and the genes were 99 to 100% identical to those of the reference K-locus*. K. pneumoniae* HS11286 and CFSAN044563 belonged to KL103, and they had a high number of capsule genes (n = 21). Furthermore, *K. pneumoniae* QS17-0029 and PIMBND2KP27 of KL50 presented the same capsular predictions (Figure S1). *A. colistiniresistens* NIPH1859 capsular predictions revealed that this isolate had KL132 with 21 capsule genes. However, this isolate was missing the allelic types and none of the genes were 100% to the primary KL reference (Figure S1).

### 3.5 Epigenomics analyses in MDR GNB

Herein, the methylome of 29 genomes are described (Table S5), of which 14, 9, three, and one isolate/s belonged to the genera *Klebsiella*, *Escherichia*, *Enterobacter*, and *Acinetobacter*, respectively. In this analysis, Type I, II, III, and IV RMS were identified in GNB isolates with carbapenem and polymyxin co-resistance. As expected, the most common system was Type II RMS, which has an important function in destroying foreign DNA ^38^. The Type II system was found in almost all isolates except four isolates (*A. colistiniresistens* NIPH1859, *E. kobei* MGH244, *E. roggenkampii* DSM 16690, and *K. pneumoniae* CFSAN044564) in which no restriction enzyme genes were found (Table S5). Type I restriction enzymes was the second most common system identified. Conversely, Type III and IV were the least common systems amongst the isolates.

The Type I system was identified in seven *E. coli* and six *K. pneumoniae* isolates (Table S5). Two *E. coli* isolates MG1655 and MCR1_NJ both honoured chromosomal Type I, II and IV systems. These two isolates had the same Type II system both comprising MTases, *M.Eco4255Dcm* and *M.EcoKII,* albeit with different motifs/recognition sequence of CCWGG and ATGCAT, respectively (Table S5). Furthermore, these isolates had methyl directed restriction enzymes Type IV system with unknown recognition sequences; except for *EcoKMcrA* in MG1655 that had a known sequence (YCGR). Only *E. coli* MG1655 and MCR-1_NJ had a complete set of chromosomal Type I system with a RE, MTase, and specificity subunit, while other *E. coli* isolates only had either MTases or restriction enzyme. However, they had the same recognition sequence. For *K. pneumonia*e isolates, KP-6884 and KL027 were the only isolates having Type I, II and III MTases (Table S5).

All *K. pneumoniae* isolates with the Type I system had both MTase and specificity subunit. *K. pneumoniae* CFSAN044571, QS17-0029 and KL027 had chromosomal MTase (*M.Kpn928I*) and specificity subunit (*S.Kpn1420I*I), while the same RMS was observed on the plasmid of *K. pneumoniae* CFSAN044563 and CFSAN044569. Furthermore, *K. pneumoniae* QS17-0029 also had a chromosomal RE (*EcoR124II*) and two more MTases; *M.EcoprrI* (chromosomal) and *M.Kpn234I* (from plasmid), which were missing in other isolates. The Type I system in *K. pneumoniae* KP-6884 was found on a plasmid hosting a MTase, *M.Kpn928I,* and a specificity subunit, *S.Kpn1420II,* that shared the same recognition sequence, ACGNNNNNNCTG.

Type II restriction enzymes were the most common restriction enzymes in bacterial. The Type II MTases included plasmid-mediated *M.Kpn34618Dcm* in *K. pneumoniae* and chromosomal *M.EcoE455Dcm* in *E. coli* isolates, sharing the same recognition site, CCWGG (Table S5). The *M.Kpn34618Dcm* was found in all *K. pneumoniae* isolates (n = 6). However, this MTase was coupled with plasmid-borne *M.EcoRII* in *K. pneumoniae* CFSAN044572 and CFSAN044573, with the same recognition site (Table S5). Furthermore, *K. pneumoniae* CFSAN044569 had two REs, viz *EarI* and *Eco128I*, and four MTases, (*M.EcoRII*, *M.Kpn34618Dcm M1.EarI* and *M2.EarI*). The *EarI, M1.EarI* and *M2.EarI* MTases shared recognition sequences, while *Eco128I*, *M.EcoRII* and *M.Kpn34618Dcm* had different motifs (Table S5). For *E. coli* isolates*, M.EcoE455Dcm* was always coupled with *M.EcoKII,* with the recognition sequence ATGCAT in the Type II system. Interestingly, the chromosomal Type II *M.EcoE455Dcm* in *E. coli* Ec36 was coupled with a RE, *Eco128I,* and two copies of *M.EcoRII* MTases (Table S5). Furthermore, the Type II system in *E. coli* WI1 was mobilised by a plasmid, with the same recognition sequences as that of the *M.EcoRII* MTases.

Types III and IV RMSs were the least common among our isolates. The Type III system was found in *K. pneumoniae* KP-6884 and QS17-0029, consisting of chromosomal *M.Kpn1420I* and *M.Kpn34618Dcm,* respectively. Type IV system was found in *E. coli* EC-129 and MRC1_NJ, consisting of plasmid MTase *EcoAPECGmrSD* and chromosomal *EcoKMrr*, respectively. The Type III MTases have different motifs, while Type IV lacks the recognition sequences (Table S5)

### 3.6 Phylogenomic analyses of included GNB

#### 3.6.1 Clonal lineages of carbapenem and colistin-resistant GNB

Isolates in this study belonged to over 100 sequence types (STs). Figure 6 presents STs that have three or more isolates. Overall, the most common clones were *K. pneumoniae* ST258 (n = 56), followed by *E. coli* ST156 (n= 35), *A. baumannii* ST191 (n = 34), ST101 (n = 29) and *K. pneumoniae* ST45 (n = 21). *A. baumannii* ST191 was only reported in Seoul, South Korea^12^. *K. pneumoniae* ST258 was the most prevalent clone in Greece (n = 50), followed by *E. coli* ST156 in China (n = 33) and *A. baumannii* ST191 in Seoul (n = 34).

**Figure 6:**
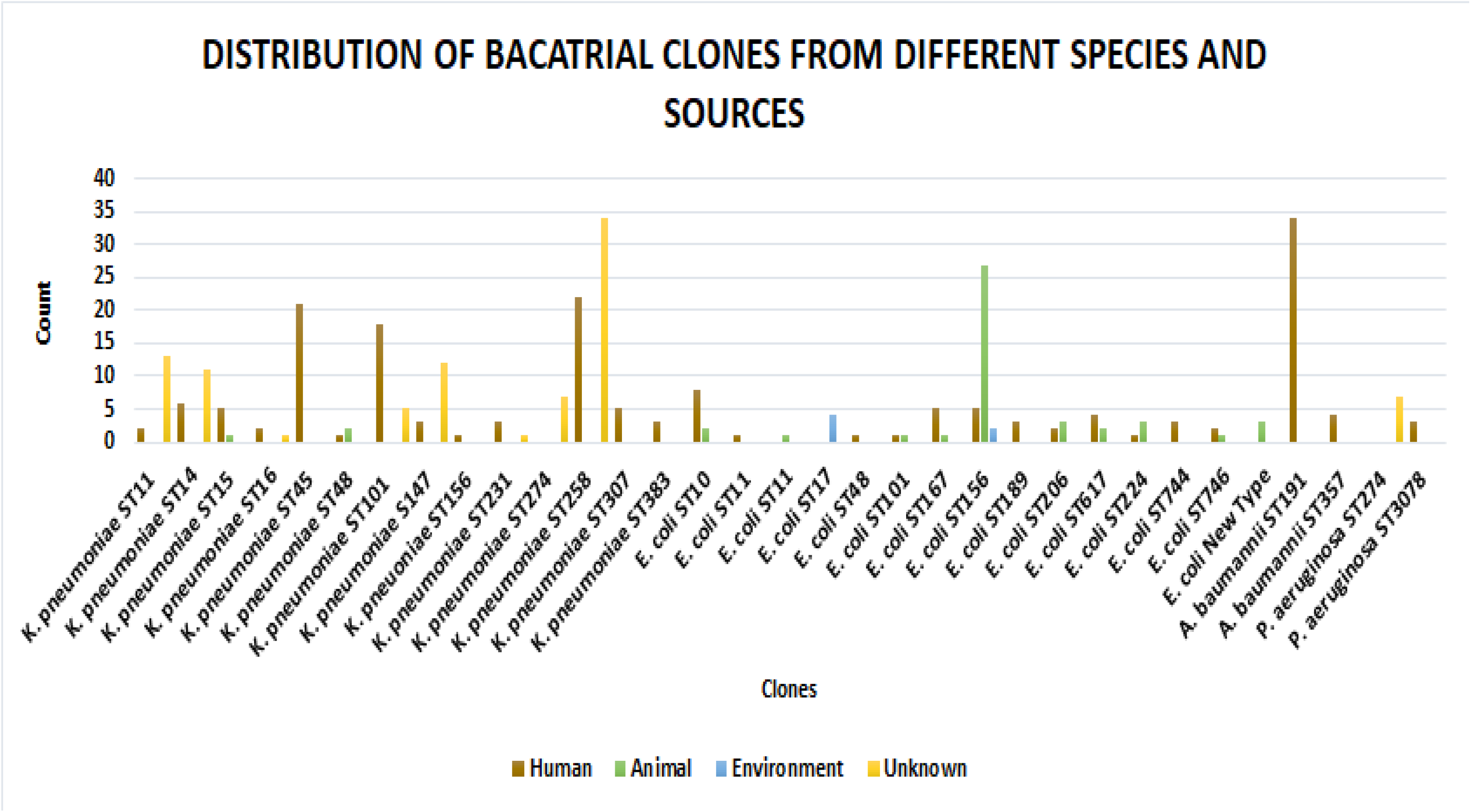
Distribution of bacterial sequence types based on country, specie, and specimen source.

Countries such as Portugal, Greece, USA, and India likewise presented diverse clonal lineages for GNB with co-resistance to carbapenem and colistin (Table S2_F). In terms of diversity across countries, clones such as *K. pneumoniae* ST15 (n = 10) was detected in seven countries, *K. pneumoniae* ST101 (n = 23) and *K. pneumoniae* ST258 (n = 17) were detected in five and four countries, respectively (Table S2_F). Furthermore, none of these clones were reported in animal samples (Table S7). Although carbapenem and colistin co-resistance was not commonly reported in African countries, Libya reported MDR *K. pneumoniae* ST101 (n = 6) and one MDR *A. baumannii* ST101. *K. pneumoniae* Kpn918 and Kpn926 of ST258 from unknown sources were found in the United States, harbouring *bla_KPC-2_* on IncFIIK plasmids. Twelve *K. pneumoniae* ST147 clones with unknown sources were reported in Greece (n = 5) and Spain (n = 7) while three isolates of the same clone were from human samples in India (n = 2) and Pakistan (n = 1) (Table S2_F). *K. pneumoniae* ST14 (n = 11) isolates with carbapenem and polymyxin co-resistance from human samples were detected in the United Arab Emirates, particularly in Dubai ^73^. *K. pneumoniae* ST14 was also reported in human samples in India (n = 6).

In contrast to *K. pneumoniae* clones, *E. coli* clones did not display high inter-country clonality (Table S2_F). *E. coli* ST156 and *E. coli* ST167 were each reported in three countries. *E. coli* ST156 was more prevalent in animal samples (n = 10) whilst *E. coli* ST167 (n=4) was found in human samples and *E. coli* WF5-38 (n=1) was from animal samples. China reported the most cases of carbapenem and colistin co-resistance in GNB, resulting in more diverse clones (Table S2_F). Plasmid replicons associated with carbapenem and colistin ARGs for these clones were also diverse. Of the 158 *E. coli* isolates that were reported in the country, most were members of *E. coli* ST156 (n = 33), followed by *E. coli* ST10 (n = 8). The remaining ST(s) only included about three isolates or less. *E. coli* ST156 was more prevalent in animal samples, accounting for 77.1% (n = 27/ 33) isolates (Table S2_F).

In addition to China, *E. coli* BJ10 of ST156 was also reported from human samples in USA. This clone harboured *mcr-1* and *bla_NDM-5_* on IncX_3_ and IncL plasmids, respectively. *K. pneumoniae* MAH-3 ST156 from humans in France also harboured *bla_OXA_* on an IncI_2_ plasmid replicon. *E. coli* ST10 clone was detected in ten *E. coli* isolates in China (n = 8) and USA (n = 2). China reported two animal and six human samples, while USA reported two isolates from humans infected with the *E. coli* ST10 clone. *E. coli* GZ2a in USA, and three *E. coli* ST10 strains in China harboured *mcr-1* and *bla_NDM-5_* respectively on an IncX_3_ and IncI_2_ plasmid. Notably, new *E. coli* clonal lineages were identified in *E. coli* WFA16004, WFA16019 and WFA16061, isolated from animals in China. These isolates harbored *mcr-1* and *blaNDM_1/4_* genes. Unfortunately, plasmid types which were associated with these ARGs are unknown ^67^.

Portugal reported a total of 25 *K. pneumoniae* ST45 isolates from human samples which harbored *mcr-1* on IncX_4_ plasmid (Table S2_A). Carbapenem and colistin co-resistance was not commonly reported in *Pseudomonas sp*., which were all detected from human samples. Ten of these were reported in Turkey i.e., *P. aeruginosa* ST235 (n = 7) and *P. aeruginosa* ST3078 (n = 3). One was reported in India, viz. *P. aeruginosa* ST235, and the last one, *P. aeruginosa* ST3357, was reported in Japan (Table S2_F).

#### 3.6.2 Phylogenetic analyses of MDR GNB

##### 3.6.2.1 Phylogenetic analyses of K. pneumoniae and E. coli isolates

Mapping the resistance dynamics, virulomes and RMS of *K. pneumoniae* isolates from human hosts against their genome-based phylogeny showed that species from the genus *Klebsiella* are highly resistant to antiobiotics (Figure 7A). However, these isolates do not possess a rich virulome and RMS. The phylogenetic tree on Figure 7A, based on *K. pneumoniae* genomes revealed three different clades, presented with different colours; Maroon (clade I), blue (clade II) and green (clade III) (Figure 7A). Clade I consisted of *K. pneumoniae* ST15 CFSAN04465 and *K. pneumoniae* ST147 CFSAN04469 from Pakistan, as well as *K. pneumoniae* ST15 MLST15 from USA. *K. pneumoniae* ST15 CFSAN04465 and MLST15 clustered together, but showed little similarities on their resistance profiles and virulomes. However, these isolates habour the same RMS, including MTase and motifs. Interestingly, *K. pneumoniae* MLST15 and *K. pneumoniae* CFSAN04469 of a different clone (*K. pneumoniae* ST147) showed significant similaries in their reistance profiles and virulence determinants. Moreover, these isolates had the same capsular predictions.

**Figure 7:**
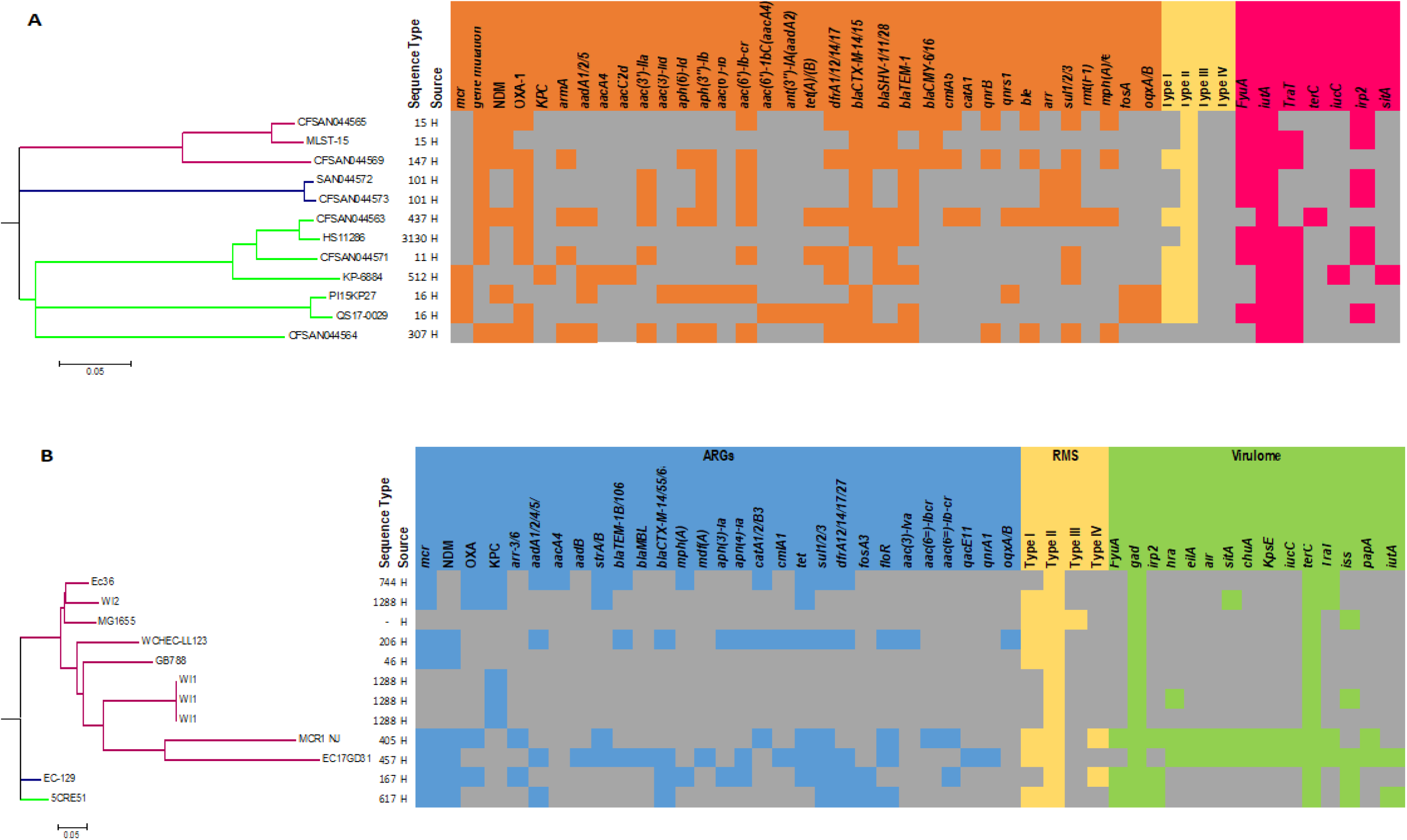
**(A):** Resistome dynamics of *K. pneumonia* isolates with carbapenem and colistin co-resistance. Orange and white blocks represents presence and absence of ARGs, respectively. **(B):** Resistome dynamics of *E. coli* isolates with carbapenem and colistin co-resistance. Red and white blocks represents presence and absence of ARGs, respectively.

*K. pneumoniae* CFSAN04472 and CFSAN04473 clustered together under clade II (Figure 7A). These isolates were isolated in Pakistan from a patient, both belonged to the *K. pneumoniae* ST101 clone and showed similar resistance profiles except that *K. pneumoniae* CFSAN04473 haboured *bla_OXA-48_* and lacked *bla_NDM-1_*. Furthermore, these isolates showed the same virulome, methylome, plasmids and capsular structures. The only difference between these isolates is that *K. pneumoniae* CFSAN04472 was isolate from blood and *K. pneumoniae* CFSAN04473 from catheter tip ^16^. Clade III was the most diverse in terms of geographical location comprising of isolates from Pakistan (n = 3), Japan (n = 1), Thailand (n = 1), Vietnam (n = 1) and Italy (n = 1). Overall, *K. pneumoniae* isolates showed different resistomes, virulomes and methylomes. *K. oneumoniae* HS11286 and CFSAN04463 belonged to different clones viz. *K. pneumoniae* ST3130 and *K. pneumoniae* ST11, respectively. These siolates showed the same RMS and virulome despite their different clonality. However, their resistance profiles were different (Figure 7A). *K. pneumoniae* CFSAN04463, CFSAN04464 and PI15KP27 belonged to different STs, but showed high sismilarities in their resistance profiles, with different virulomes and methylomes (Figure 7A).

Figure 7B depicts the evolution and resistome dynamics, virulomes and RMS of *E. coli* isolates based on genome sequences. Isolates were all from human samples. Overall, grey blocks on the *E. coli* resistome analyses is more dominant than the red blocks, meaning the *E.coli* isolates are not as highly resistant as the *K. pneumoniae* isolates (Figure 7A & B). However, there is an interesting exception for three *E. coli* isolates, viz. EC-129, EC17GD31 and WCHEC-LL-123, which harboured more ARGs besides those of carbapenems, colistin and tigecycline (Figure 7B). These species were all from human samples and were isolated from Japan (EC-129), China (WCHEC-LL-123), and Vietnam (EC17GD31). Interestingly, *E. coli* isolates revealed a more diverse viruolome profiles as compared to those of *K. pneumoniae* MDR isolates (Figure 7B).

The genome phylogeny of *E.coli* also showed three distinct clades shown by Maroon (clade I), blue (clade II) and green (clade III). Clade I comprises of isolates from China (n = 2), France (n = 2), Portugal (n = 1), USA (n = 1) and Vietnam (n = 1). Clade II and III consist of one isolate each. *E. coli* EC129 and 5CRE51of clade II were respectively isolated from Japan and Taiwan. Although most of the *E.coli* isolates clustered under clade I, they showed very distinct methylome, virulome, plasmids and resistance profiles. All isolates belonged to a distinct clone, except for *E. coli* ST1288 WI1 and WI2 from France (Figure 7B). Furthermore, these two isolates had similar virulence factors. *E. coli* WI2 of ST1288 and *E. coli* Ec36 of ST644 had the same virulome and simlar resistance profiles. *E. coli* MCR1_NJ (ST405) and *E. coli* EC17GD31 (ST457) revealed similar resistance profiles and RMS, where MCR1_NJ had a Type IV restriction system that was rare in the included isolates. Furthermore, the two isolates showed similar virulomes, except that *iss* and *iut*A were missing in *E. coli* MCR1_NJ (Figure 7B).

##### 3.6.2.2 Plasmids evolutionary analyses

Figure 8A shows phylogenetic analysis of plasmids associated with carbapenemases, while Figure 8B shows that of plasmids associated with polymyxin resistance. Both phylogenies revealed six distinct clades. On Figure 8A, clade I consisted of four *E. coli* plasmids from human samples which belonged to different STs and harboured different plasmids. However, two of these *E. coli* isolates did not have plasmids replicons (Figure 8A). Clade II showed a bootstrap of 100 and consisted of *E. coli* and *K. pneumoniae* plasmids. It is worth noting that although strains in clade II belonged to different STs, they harbour IncX_3_ plasmids, except for *K. pneumoniae* DHQP1605752 that had A/C plasmid. Furthermore, the lower subgroup of this clade consisted of four *E. coli* isolates (P768MEM-011, P788A-032, P785MEM and P744T) from animal samples which clustered tightly (Figure 8A).

**Figure 8:**
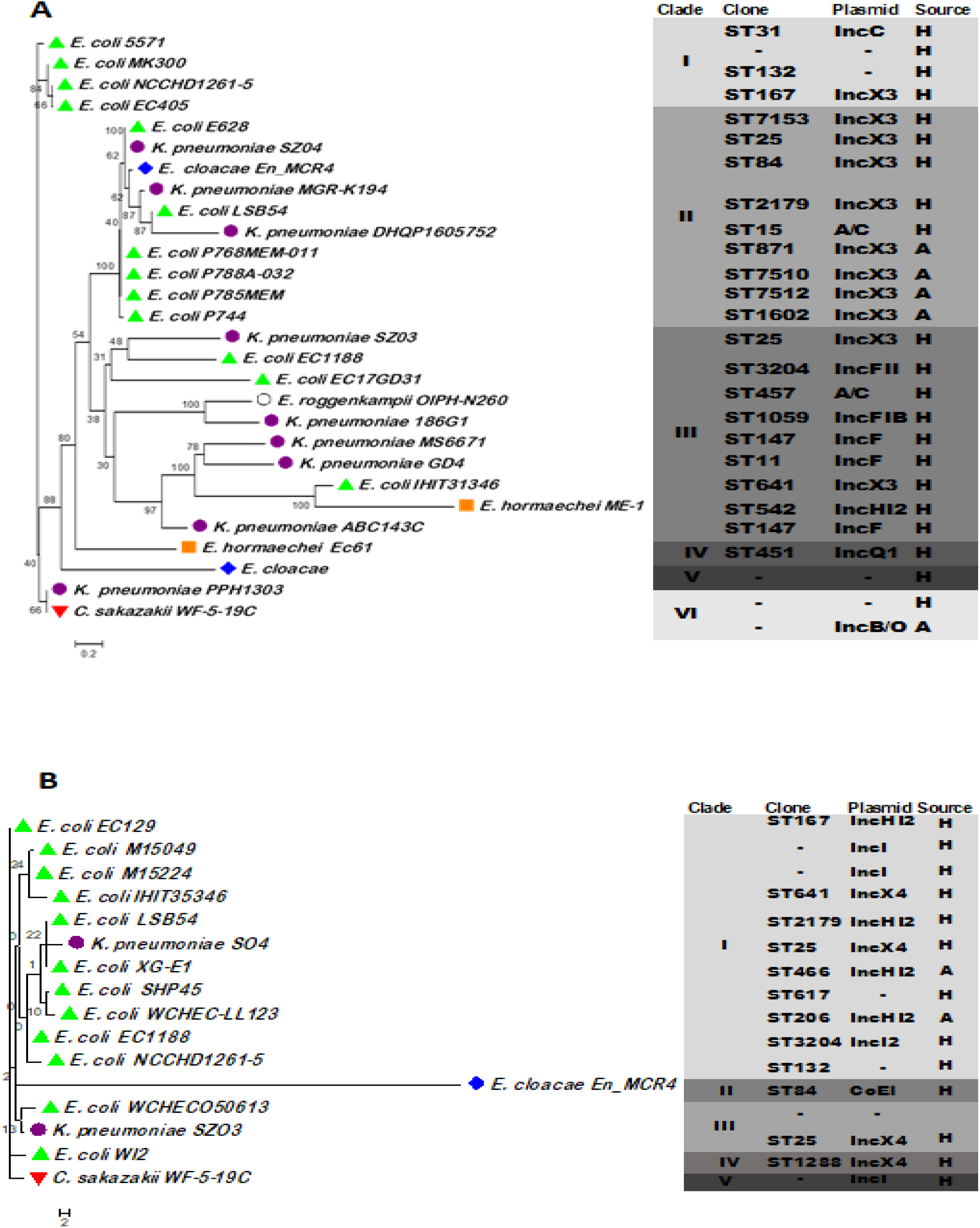
Evolutionary phylogenetics of plasmids harbouring carbapenem and polymyxin resistance determinants. **(A)** Plasmid evolutionary analyses of plasmids replicons associated with carbapenemases. **(B)** Plasmid evolutionary analyses of plasmids replicons associated with colistin ARG, *mcr-1*. *E. coli, K. pneumoniae, E. cloacae, E. roggenkampii, E. hormaechei* and *C. sakazakii* isolates are shown by green triangle, purple circle, blue diamond shape, clear circle orange square and red triangle, respectively.

Clade III was the most diverse clade consisting of carbapenemase-associated plasmids from *K. pneumoniae* (n = 5), *E. coli* (n = 2), *E. roggenkampii* (n = 1) and *E. hormaechei* (n = 1). These plasmids belonged to different STs and were isolated from different locations. However, plasmids in clade III were all from human samples. *K. pneumoniae* GD4 and SZ03 harboured IncX_3_ plasmid, which was more prevalent in clade II. However, this plasmid carried *bla_KPC_* and *bla_NDM_* gene in *K. pneumoniae* GD4 and SZ03, respectively. Clades IV and V were singletons consisting of plasmids from *E. hormaechei* Ec61 (clade IV) and *E. cloacae* (clade V) from human samples. Lastly, clade VI had one isolate from human origin and one from animals with unknown sequence types (Figure 8A).

Figure 8B shows phylogeny, resistance profiles, RMS and virulome profiles of plasmids that mobilize the *mcr-1* gene. In contrast with carbapenemase-borne plasmid-based phylogeny, this phylogeny comprised of a more diverse clonality. All plasmids in clade I belong to their unique clones. *K. pneumoniae* SZO4 and *K. pneumoniae* SZO3 in clade III plasmids (IncX_4_) belong to the same clone ST25 (Figure 8B). IncHI*2* plasmids were found in four *E. coli* isolates of different clones. Two of these plasmids were detected in two *E. coli* WCHEC-LL123 and XG-E1 isolated from animal samples. However, these isolates belong to *E. coli* ST206 and *E. coli* ST466, respectively (Figure 8B).

## 4. Discussion

Increasing antimicrobial resistance in GNB continue to challenge public health globally. Carbapenem and polymyxin co-resistance is even more worrying as these antibiotics are “last-line” treatment therapy for GNB infections. Here, we present in-depth genomic epidemiology, epigenomics, virulome and mobilome analysis of MDR GNB using a One Heath concept, globally.

Various countries evidently showed that carbapenem and polymyxin co-resistance disseminate among humans, animals, and the environment. In addition to high numbers of carbapenem and colistin co-resistant isolates, China presented the most diverse specimen from various sources, as well as multiple clones (Table S2_A). The observed outcomes could be due to the over-prescription of antibiotics in health facilities and overuse of antimicrobials for animal growth and wellness ^74^.

Although GNB in humans have been the most common reservoirs for antibacterial resistance genes, this trend is now increasing in animals, particularly in swine and poultry in developing countries ^75^. Thus, co-resistance to carbapenem and colistin in GNB have been outspread globally, affecting both human health facilities and veterinary medicine ^1, 76^. Countries such as China, Japan, USA, and Italy have reported carbapenem and colistin co-resistance in both human and animals’ samples. Carbapenem and colistin co-resistance in bacteria from animals is of great concern in Asian countries, particularly in China ^32, 77^. Previous studies showed that antibiotic resistance in livestock drastically increased in the past two decades in the North-eastern parts of China and India ^75^. This is due to the increasing demands of meat in developing countries. Such countries frequently use antibiotics to boost animal wellbeing and growth to elevate meat production ^75, 78^. Even though Asia and Europe reported high numbers of carbapenem and colistin co-resistance, European countries reported less numbers from animal samples, because Europe and the United States prohibited the use of antibiotics as growth promoters in animals ^78^. Environmental isolates were only reported in two countries, China, and Germany. These were from farms, slaughterhouse, and sewage systems ^67, 71, 79^ which suggest that carbapenem and colistin co-resistance may not be common in environmental samples as it is in isolates from animal and human origin.

*K. pneumoniae* are commonly known for nosocomial infections, which are often untreatable and fatal ^3, 22^. As expected, our results revealed that *K. pneumoniae* isolates were commonly reported for carbapenem and colistin co-resistance globally (n = 535) (Figure 2A). Other GNB species such as *E. coli*, *P. aeruginosa* and *A. baumannii* are also implicated in nosocomial infections ^2, 4^. Additionally, Enterobacter spp. such as *E. cloacae* and *E. hormaechei* have recently been identified as important pathogens in health facilities ^80^. *E. cloacae* isolates are naturally resistant to many antibiotics including first-generation cephalosporins, ampicillin, and amoxicillin, which makes them multi-antibiotic resistant ^80^. This explains their rich resistance profiles found alongside carbapenem and colistin co-resistance (Figure 4A &B).

Furthermore, these isolates harboured the major plasmid replicons reported in this study viz., IncI, IncX IncF, and IncHI (Figure 5A &B), which may play a role in the easy transmission of ARGs. *P. aeruginosa* and *A. baumannii* isolates revealed low ARG profiles and/or lack of plasmids, virulence genes, RMS and capsular polysaccharides. This suggests that some *P. aeruginosa* and *A. baumannii* isolates are not MDR and their infections may still be treated successfully with antibiotics. Our results revealed that for all *P. aeruginosa* and *A. baumannii* with carbapenem and colistin co-resistance, there were no plasmid-borne *mcr* genes, but only chromosomal resistance mechanisms were identified (Table S2_A). This corroborates the lack of plasmids in these isolates.

The reported isolates belong to various clones. The most common clones with carbapenem and colistin co-resistance were *E. coli* ST156 in China, *K. pneumoniae* ST258 in Greece and *A. baumannii* ST191 in Korea (Table S2_F). The international *K. pneumoniae* ST258 was reported in more than one country, indicating its easy transmissibility and wider distribution. This is an international clone that is continually being reported globally ^22^. This means core resistance genes will be easily disseminated between bacteria, which makes the fight against antibiotic resistance in GNB, particularly *K. pneumoniae* of ST285, almost impossible to win. Furthermore, having this promiscuous clone now involved in co-resistance to the two most important antibiotics calls for the development of effective treatment therapies or even new drugs.

*E. coli* isolates belonged to multiple STs, but *E. coli* ST156 was the most common (Table S2_F).This epidemic clone is related to the spread of *mcr-1*, NDM and CTX-M ^81^, which is also supported by our results where *E. coli* ST156 isolates from human and animals were found to carry *mcr-1* and *bla_NDM_* ARGs in China and USA (Table S2_A).Furthermore, new *E. coli* STs from animals have been reported in China ^62^. The emergence of new clones and high numbers of animal cases suggests that GNB are continually evolving and the crisis of carbapenem and colistin co-resistance is undoubtedly growing. This also suggest that there may be an increasing use of antibiotics in animals for commercial and economic benefits*. A. baumannii* ST191 was another common clone in our analyses. It was prevalent in *K. Pneumoniae* isolates and was only identified in Korea ^12^. This clone carried OXA-23 along with the disrupted *mgrB* gene for carbapenem and colistin resistance, respectively (Table S2_A).

Many studies suggested combination therapies as treatment alternatives due to increasing ESBLs, carbapenemases, and now polymyxin resistance in GNB ^7, 13^. Unfortunately, our results revealed that isolates with carbapenem and colistin co-resistance also developed resistance against other antibiotics including tigecycline, aminoglycosides, chloramphenicol, and sulfamethoxazole (Figure 4), therefore challenging combination therapies as treatments. Resistance to various antibiotics, in addition to the rich virulence factors, methylome and capsular polysaccharides (in *K. pneumoniae*) explains the success of *K. pneumoniae* to bypass the host’s immune defences, making their infections more challenging to treat/manage^3^. NDM, OXA-48 and KPC were previously reported as the most widely distributed carbapenemases, with KPC being the most prevalent in *K. pneumoniae* ^3^.

However, our results revealed that NDM and OXA-48 were predominantly associated with polymyxin resistance globally. An increase in the production of OXA-48 in *K. pneumoniae* isolates was reported in 2019 ^82^, which was also observed in our analyses where OXA-48 was the most most prevalent carbapenemase, followed by NDM (Figure 3B). Similar to other isolates reported in India, Table S2_A showed that *K. pneumoniae* isolates in United Arab Emirates (Dubai) also produced OXA-48 and NDM. However, the polymyxin resistance mechanism for these isolates were not detected. KPC was previously reported as the most prevalent carbapenemases in *K. pneumoniae* ^4^. However, in this study OXA-48 was detected more than KPC globally. OXA-48 was predominant in *K. pneumoniae* isolates from India. OXA-48 was reported to have lower activity against carbapenem but, showed increased activity when it co-exists with ESBLs ^19^. Our result showed that OXA-48 was coupled with ESBLs in most isolates (Figure 4), which supports their resistance to carbapenems.

Colistin resistance was through plasmid mediated *mcr-1* and/or chromosomal mutation. Furthermore, Table S2_Ashows that *mcr-1* is carried by different plasmids, suggesting a rapid and high dissemination of this gene, thus increasing colistin resistance in bacteria. Unfortunately, these plasmids have spread to carbapenemase-producing isolates, resulting in MDR bacteria. Resistance through *crrB* gene was the least common mechanism for polymxin resistance. This mechanism has recently been reported as another pathway to polymyxin resistance in GNB ^83^. This suggests that bacteria are continuously evolving, resulting in the emergence of new resistance mechanisms.

DNA methylation play a crucial role in the regulation of gene expression and it is part of bacterial defence systems^37^. Bacteria implement systems that are very crucial in destroying foreign DNA material through recognition sequences ^35, 37^. Most *K. pneumoniae* isolates share the same methylation signatures as *E. coli* isolates, suggesting that DNA & MGEs can be easily exchanged between the two species, thus giving them an advantage in acquiring resistance and virulence to more antibiotics and hosts, respectively. Likewise, *mcr’*s ability to easily disseminate as compared to other mechanisms is hinged on its association with MGEs. This suggest that plasmids that carry the *mcr-1* gene may have the same recognition sequences as the host, which makes their transmission between bacteria a success.

The MGEs that carry carbapenem and colistin ARGs are also associated with a diversity of virulence genes in MDR GNB. Bacteria also produce polysaccharide capsules for their protection, which are one of the most important virulence factors in pathogens ^38^. Bacterial virulence genes/factors enable them to bypass the hosts’ defence systems ^84^. MGEs, RMSs, virulence genes and capsules all contribute to AMR in GNB. The most common mobile elements were plasmids, followed by insertion sequence (IS), transposon (Tnp) and class I integrons (Table S3_A). IncX plasmids were previously identified as the main carrier of carbapenem and colistin ARGs among GNB ^12, 66^. In line with this, our study revealed that IncX was the most common MGE in 43 countries, carrying carbapenemases and *mcr-1* gene (Table S3_B). China, with the highest reported numbers of carbapenem and colistin co-resistant isolates (n = 171), presented diverse plasmids from various sources (Figure 2C). Surprisingly, USA (n = 18), Japan (n = 15), and Vietnam (n = 6) were among countries with the most diverse plasmid replicons despite their low rates of carbapenem and colistin co-resistance. However, this was expected for Japan as it presented a diverse species distribution like China (Figure 2A). The diverse plasmid repertoire in USA and Vietnam could suggest that there are chances of increasing rates of plasmid borne ARGs within these countries. The insertion sequence IS903B was the most common and prevalent in *K. pneumoniae* isolates (p = 0.362). This IS was said to be one of the main elements that facilitated *mcr-8*’s dissemination in GNB ^85^. The co-occurrence of the *mcr-*8 with NDM was previously documented in *K. pneumoniae* ^85^.

Colistin resistance has long been occurring through chromosomal mutations. However, the emergence of plasmid-mediated *mcr* genes has put more strain to public health because such genes are easily transmitted than chromosomal resistance mechanisms ^24^. As previously stated ^86^, our study revealed that *mcr* co-occurrence with carbapenemases were commonly identified in *E. coli* and showed low detections in *K. pneumoniae*. Further, *mcr* was not identified in any *A. baumannii* and *P. aeruginosa* isolates. This evidently explains the low rates of colistin resistance in *A. baumaniii* and *P. aeruginosa* isolates.

We showed that carbapenem and colistin co-resistant *K. pneumoniae* isolates from human samples clustered under three distinct clades. Likewise, carbapenem and colistin co-resistant *E. coli* isolates resulted in three clades, with clade I comprising of more isolates while clades II and III comprised of single isolates (Figure 7A & B). In addition to carbapenem and colistin ARGs, *K. pneumoniae* and *E. coli* showed a rich resistance profile, confirming their potential as carbapenemase-producing and polymyxin resistant MDR bacteria. *K. pneumoniae* ST101 isolates from the same location showed the same plasmids, virulence genes, methylome and capsules. However, their resistance profiles were not entirely identical (Figure 7A). Contrarily, *K. pneumoniae* ST15 isolates of clade I of the *K. pneumoniae* phylogeny had differences in their resistance profiles and virulomes. This suggests that bacterial resistomes and virulence determinants are independent of their sequence types, which could be due to the presence of different plasmids in the same clones.

Furthermore, these isolates showed the same RMS, capsular polysaccharides, and were isolated from different countries. Additionally, this proves that their RMS are also independent from their location of isolation. Having the same RMS allows them to easily transmit genetic material, given that they have the same recognition sequences. Likewise, phylogenetic analyses based on *E. coli* genomes also support the finding that bacterial virulomes and RMS are independent of clonality (STs), source, and geographical location (Figure 7B). These analyses revealed that genome sequences provide higher resolution than STs, and thus show a better relationship between isolates than STs. Therefore, whole genome sequences should be used to type or distinguish clones in epidemiological studies and outbreaks.

Phylogenetic analyses based on plasmids demonstrated that with a few exceptions, the same plasmids carrying the same carbapenemase (Figure 8A) and colistin (Figure 8B) ARGs, mostly cluster together regardless of their sequence type and source of isolation. The observed patterns of clustering suggest that plasmids associated with the same ARGs may be genetically related. This is supported by our results that show IncX plasmids as the most common plasmid to host both carbapenemase and polymyxin resistance genes. Hence, the presence of same plasmids in different species indicates that plasmids are constantly transmitted among GNB.

### Conclusion, recommendations, limitations, & future perspectives

One of the major global crises currently facing human and veterinary medicine is increasing antibiotics resistance in bacteria. High demands and over prescription of antibiotics have resulted in an increasing burden of bacterial antibiotic resistance globally. An even more worrying problem is the emergence of bacterial resistance against last-resort antibiotics such as carbapenems and polymyxins. This was commonly reported in GNB isolates from animal, human, and environmental sources. Although carbapenem and polymyxin co-resistance was common in *K. pneumoniae*, a stronger association existed in *E. coli* isolates from animal samples. Countries with high demand for meat production viz., China, India, and Italy, reported more cases of carbapenem and polymyxin resistance in animals.

Continuous dissemination carbapenem and polymyxin ARGs is supported by various MGEs, RMS systems and rich virulence determinants. Reported MGEs were integrons, insertion sequences, transposons, and plasmids. These were associated with plasmid-borne *mcr-1* that confers resistance to colistin and with plasmid-borne carbapenemases such as OXA, NDM, KPC, IMP, GES-5, and VIM. A very high number of clones were reported globally for GNB with carbapenem and polymyxin co-resistance. The most common clones were *E. coli* ST156 in China, *K. pneumoniae* ST258 in Greece and *A. baumannii* ST191 in Korea. Evidently, these clones each showed their unique resistance profiles, RMS, capsules, virulomes and MGEs, supporting their independent evolutionary trajectories. A new *E. coli* clone was reported in China from animal samples. Such clones need to be studies further.

The lack of available genome and/or plasmid accession numbers from published articles limited our bioinformatics studies. Our study showed that there is an urgent need to institute stricter regulations on antibiotic stewardship, prescription, and sales to prevent a return to the pre-antibiotic era as the global dissemination of these MDR strains can take many lives. Further, new antibiotics or new treatment therapies that can combat antimicrobial resistance are urgently needed. Furthermore, more genomic investigations are needed to map current resistance mechanisms and trends globally to facilitate evidence-based clinical and epidemiological decisions in future.

## Data Availability

All data for this work are included in supplemental files

## Availability of material and data

Supplementary data is available and included as Tables and Figures in this manuscript.

## Acknowledgements

None

## Transparency declaration

The authors declare no conflict of interest.

## Author contributions

Study design and supervision (Dr. J.O.S); Bioinformatic/statistical analyses (W.T.R & Dr. J.O.S); Write-up (W.T.R & J.O.S); Editing, revision, and formatting (Dr. J.O.S).

## Supplementary Figures

**Figure S1:**
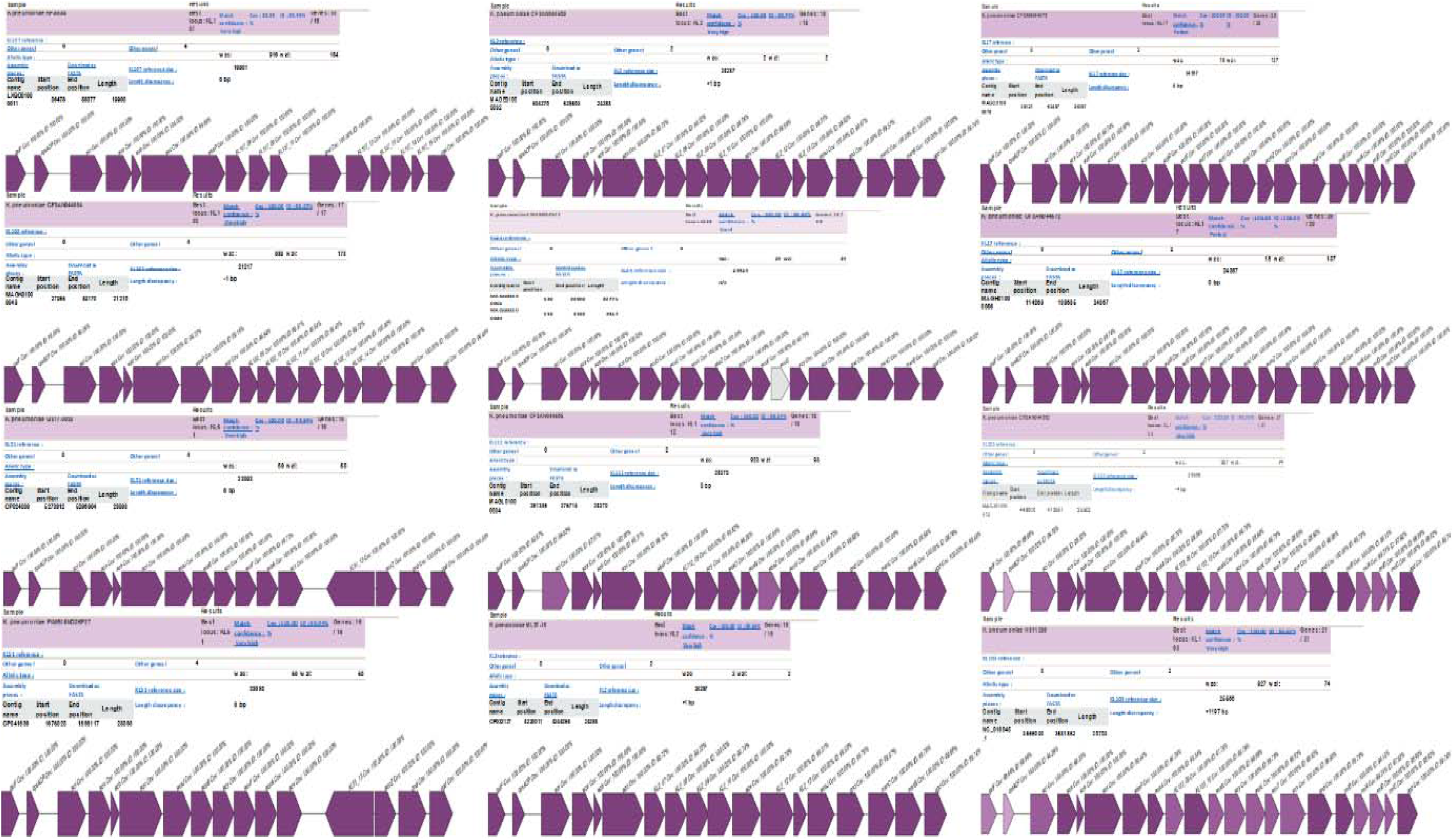
Capsular polysaccharide predictions of carbapenem and colistin resistant GNB based on the K-locus of *K. pneumoniae*.

## Supplementary Tables

**Table S1. Excluded and included articles. (A)**: Data showing excluded papers and (**B)**: shows papers included for meta-analyses.

**Table S2. Dataset of extracted data from included articles. (A)**: Data showing geographical location, sample source, species, clones, genetic environment, antibiotic resistant genes (ARGs), mobile genetic elements (MGEs) and plasmids replicons/incompatibility (Inc) of GNB isolates which are co-resistant to carbapenem and colistin, **(B):** species distributions per country, **(C):** specimen source in different countries, **(D):** ARG distributions in different countries, **(E):** ARG distributions in different species, **(F)** Sequence types for isolates with carbapenem and colistin co-resistance globally based on country of isolation, specimen source and species type.

**Table S3. Dataset of genomic analysis of downloaded genomes. (A):** Data showing the MGEs that are responsible for the transmissions of carbapenem and colistin resistance among GNB, **(B):** global distribution of plasmid replicons associated with carbapenemases and colistin ARGs from included articles and **(C):** Plasmid distributions in different species.

**Table S4: Analysis of virulence factors (virulome) in carbapenem and colistin resistant isolates from various countries.**

**Table S5:** Methylome data of carbapenem and colistin resistant isolates showing Type I, II, III and IV restriction modification systems in different isolates.

